# Transitions in childbirth care provision: Understanding the rapid rise in institutional delivery in 21 countries of sub-Saharan Africa and the implications for future strategies

**DOI:** 10.1101/2025.10.20.25338390

**Authors:** Andrea K. Blanchard, Amanuel Abajobir, Martin K. Mutua, Fernando C. Wehrmeister, Anne W. Njeri, Godfrey Adero, Dieneba Aidara, Arsène B. Sandie, Luís Paulo Vidaletti Ruas, Cauane Blumenberg, Cheikh M. Faye, Ties Boerma, The Countdown to 2030 MNH Study Collaboration

**Affiliations:** Institute for Global Public Health, University of Manitoba, R070-771 McDermot Avenue, Winnipeg MB R3E 0T6, Canada; African Population and Health Research Center, Kitisuru, Manga Close, Kirawa Road, Nairobi, Kenya, P.O. Box 10787-00100; International Center for Equity in Health, Federal University of Pelotas, R. Mal. Deodoro, Pelotas - RS, Brazil 96020-220; African Population and Health Research Center, West Africa Regional Office (WARO), 4e étage, Immeuble Sourok 3, 10083 Sacré coeur 3 VDN, Dakar, Senegal; Muso Health, United States, 3254 19th Street, 2nd Floor, San Francisco CA 94110, USA; causale consultoria, Pelotas – RS, Brazil 96090-840

**Keywords:** maternal health, newborn health, childbirth care, institutional delivery, healthcare quality, health equity, sub-Saharan Africa

## Abstract

**Background:** In sub-Saharan Africa (SSA), institutional births have risen rapidly but mortality has remained high. We examined whether there have been increasing births at hospitals, with skilled attendance, and emergency capacity as indicators of comprehensive, higher-quality childbirth services in 21 SSA countries over the last two decades.

**Methods:** We analysed national household surveys between 2001 and 2022 to examine population trends in birth place (hospital or lower-level), attendant, and Caesarean Section (CS) rates by wealth quintile, and routine health facility data for 2022 on volumes of live births and CS by facility level. Countries were classified based on recent institutional delivery coverage (<65%, 65-85%, >85%), to assess patterns of change and future directions in line with a maternal and neonatal mortality transition model.

**Results:** Institutional delivery increases were primarily driven by lower-level facilities, which had low birth volumes and limited CS capacity. Yet countries that reached high delivery coverage saw greater gains in hospital births, attendance by doctors, and CS rates among the poorest. As national coverage rose, more deliveries were conducted at higher-volume CS-capable hospitals. Low population CS rates among the poorest persisted everywhere.

**Conclusion:** Major increases in institutional deliveries have not sufficiently translated into equitable access to comprehensive, life-saving childbirth care in 21 countries of SSA. Shifts towards hospital deliveries in countries that reached high coverage, consistent with the transition model, can provide guidance to those with lower coverage (<85%). Contextualizing strategies to equitably provide high-quality childbirth care will be transformative for women’s and newborn’s health in SSA.

## 1. Background

Despite declines in the last two decades, rates of maternal, stillbirth, and neonatal mortality in low- and middle-income countries (LMICs) remain high and progress has been uneven (1,2). Comprehensive maternal and newborn health (MNH) care is paramount and requires continual improvements to accelerate mortality declines (3–5). The maternal mortality, stillbirth, and neonatal mortality transition model captures five phases of mortality reduction (5). Countries in the last two phases, which have addressed most preventable mortality, are typically characterized by near-universal institutional delivery, with a predominance of births at hospitals rather than lower-level (LL) (or non-hospital) health facilities, and Caesarean section (CS) rates among the poorest groups that meet the need for emergency care.

In sub-Saharan Africa (SSA), there has been a dramatic increase in institutional births in recent years. Evidence suggests that this surge has primarily occurred at LL facilities such as clinics and health posts (4,5). These facilities have less resources than hospitals to meet the essential standards of life-saving emergency obstetric and neonatal care (EmONC), and have challenges with timely referrals in emergencies, which undermine the ability to ensure safe outcomes (3,4,6–8). This raises important questions on whether the increases in service utilization in the region have been accompanied by achieving progress towards comprehensive emergency obstetric and neonatal care (CEmONC) (4,5,9,10). Wider debates remain over the ideal organization of services to provide childbirth care that optimizes resource allocation, with some recommendations for universal hospital births attended by doctors or specialists, though there is no global consensus to date (11,12). While studies have shown trends, inequalities, and related factors for institutional delivery and CS rates across SSA (13–19), few have considered the facility level (e.g. hospital versus LL facilities) in relation to wealth-based inequalities or volumes of births and CS assisted deliveries across the region (3,6). Multi-country evidence is therefore needed to better understand patterns, trends, and inequalities in where births occur and with what emergency capabilities to guide future MNH strategies.

To address these gaps, a multi-country study was conducted to describe trends in childbirth care provision over the past two decades in 21 SSA countries, focusing on two related questions. First, to what extent has the dramatic increase in institutional deliveries been accompanied by a shift toward hospital births, more skilled attendance, and increases in CS rates, particularly among the poorest populations? Second, has the rise in institutional deliveries resulted in a higher proportion of births in higher-volume facilities and facilities with high CS volumes, which may be associated with higher quality of care? The paper presents and discusses the findings in light of the mortality transition model benchmarks, related literature and countries’ MNH-related policies, and offers recommendations for future strategies to ensure equitable access and quality of childbirth care in the region.

## 2. Methods

### 2.1 Design and data sources

The study was conducted by the Countdown to 2030 for Women’s, Children’s, and Adolescents’ Health initiative, which involved global partners and collaborators from 21 SSA country teams who were interested and provided data to participate. These country teams include analysts from academic institutions and national Ministries of Health aiming to strengthen national capacity for evidence generation, track progress, and enhance accountability. This study analyzed Demographic and Health Surveys (DHS) and Multiple Indicator Cluster Surveys (MICS), which are representative at the population level (20,21). Survey years were chosen to be as similar as possible among countries’ available datasets within the study period of 2001-2022 to aid trend comparisons (Table A.1). The study considered DHS data from women of reproductive age (15-49 years) with at least one live birth in the last three years for DHS and the last two years for MICs surveys. In addition, the study analyzed routine health facility data from each country’s Health Management Information System (HMIS) from January to December 2022. Data are reported by health facilities to district offices monthly using standard reporting forms that may differ between countries, and then usually entered into digital form using the web-based platform, District Health Information System software (DHIS2). Initial data quality checks were conducted by the Ministries of Health to correct issues.

### 2.2 Indicators

Using the surveys, we analyzed trends in the proportion of live births by place of delivery and type of birth attendant. Place of delivery was categorized as: 1) hospital, 2) LL health facility, and 3) home or other non-facility settings. Birth attendants were categorized as: 1) doctor, 2) nurse or midwife, 3) other skilled professional (considered as a skilled attendant), or 4) other non-skilled attendant or no one. Original survey categories were combined into the above categories in consultation with the country teams to best reflect their respective health systems’ organizational structure (Table A.2). We then assessed the proportion of births by place and attendant together. We also examined trends in population CS rates, defined as the proportion of live births that were delivered by a CS. Institutional CS rates were also computed, as the population CS rate divided by population institutional delivery rate. Higher values suggest that a higher proportion of institutional deliveries are being managed with CS, highlighting better access to emergency services – a marker of quality of care. The assessment of met need for CS is focused on the extent to which the poorest women receive the intervention, as non-medically indicated CS is uncommon among the poorest in the region (18,22). We stratified these indicators by wealth quintile using asset indices calculated based on asset ownership and household characteristics, such that the poorest are in quintile one and the richest in quintile five. Wealth index scores are computed by the original survey teams for each household using Principal Component Analyses (PCA), then divided into five quintiles (poorest 20% as Q1 to wealthiest 20% as Q5).

The HMIS/DHIS2 contains data on all services offered at the health facility, including reproductive, maternal, newborn, child, and adolescent health (RMNCAH) services. DHIS2 data were extracted monthly at the facility level for the last complete year (2022). MNH records were extracted containing the monthly totals for all births, live births, deliveries, assisted vaginal deliveries, CS, and stillbirths. The present study focuses on live births and CS among live births. Countries used master facility data to extract details about type, ownership and facility level in each country. Given the different service organization in each country, all facility types were classified as either hospitals or non-hospitals (Table A.3).

### 2.3 Data management and quality

For survey data, variables were compared to the standardized institutional delivery and skilled birth attendance indicators from the International Center for Equity in Health (ICEH) database to evaluate consistency (23). Absolute differences larger than two percentage points (pp) were individually assessed and addressed. Additionally, face validity checks were performed for each indicator to align with expected patterns and contexts.

In preparing the routine facility data, the first step was to merge the administrative and MNH facility records to incorporate information on the region, district, level, type and ownership of the health facilities. After merging, each country team cross-verified the number of facilities that reported any births that matched with the administrative data (to reflect facilities in the database merging with the facility master list). Facilities that merged successfully were used for analysis (Table A.4). The median merging rate across countries was 98.4%. Data quality checks and adjustments were performed before analysis, in line with standard Countdown quality checks, adapted for the use of monthly facility reports rather than aggregated to the district level (24,25).

### 2.4 Statistical analysis

Survey data were analyzed accounting for sampling design and weights within each country’s datasets. Descriptive analyses were conducted for each country by computing frequencies, interquartile ranges, and confidence intervals. Using countries’ recent survey estimates for institutional delivery coverage, we created three groups: lowest (<65%), medium (65-85%), and highest (>85%), and compared their medians for more the specific delivery care indicators at each time point and absolute changes between them. The median year of the countries’ early surveys was 2005, and the median year of their recent surveys was 2019.

According to the 2023 UN estimates for maternal mortality ratio (MMR), stillbirth rate (SBR), and neonatal mortality rate (NMR) (1,2), almost all study countries are now in transition Phase II or III (Nigeria remained in Phase I only for MMR) (Table B.1). The transition model also includes benchmark coverage levels that are empirically associated with each mortality phase, including deliveries at hospitals and CS rates among the poorest (Table B.2) (5). Therefore, we compared the study countries’ recent coverage levels with the typical values for all countries in Phase IV as the next lower mortality phase – defined by MMR <100 per 100,000 and SBR plus NMR <30 per 1,000 births – as a reference for future improvement in delivery coverage measures needed to further lower mortality levels (5).

HMIS/DHIS2 data were analyzed by summarizing total and median volumes of live births and CS by facility level (hospitals versus non-hospitals). Key descriptive statistics, including annual totals, medians, and interquartile ranges (IQR), were calculated. We also assessed the proportion of live births at hospitals and non-hospitals with lower and higher volumes of live births and CS. For both indicators, we used an arbitrary cut-off of above or below 183 in a year (or one every other day) at non-hospitals, and above or below 500 in a year (over one per day) at hospitals, respectively. While no common definitions exist to our knowledge, these cut-offs accommodated all countries’ IQR to allow comparisons, and align with prior analyses using interpretable cut-offs based on days and weeks in the year (3,6). Earlier evidence also suggests that having at least 500 births in a year, especially those with CS capacity, was associated with significantly higher quality of maternity care than lower volumes (6). All analyses were carried out in STATA 18.0 and Excel.

## 3. Results

### 3.1 Has the rapid rise in institutional deliveries in 21 SSA countries been accompanied by increasing births at hospitals, with skilled attendants, and CS rates among the poorest (DHS/MICS)?

Between 2005 and 2019 (median baseline and endline years), institutional delivery rates increased substantially across 21 SSA countries, from a median of 43.8% (IQR 32.8-54.1) to 80.3% (IQR 66.5-85.4) across countries, respectively (Figure 1). The most substantial absolute increases were observed in countries that achieved the highest coverage (≥85%) in the recent surveys, followed by those in the middle (65-85%) and lowest (<65%) coverage groups, with median increases of 47.4, 32.7, and 24.1 pp, respectively. Institutional delivery coverage in the highest coverage group is already in the range of typical values for countries in the transition model’s Phase IV (with median 97%, IQR of 88-99%), but countries in the middle and especially the lower coverage group have room to improve.

**Figure 1:**
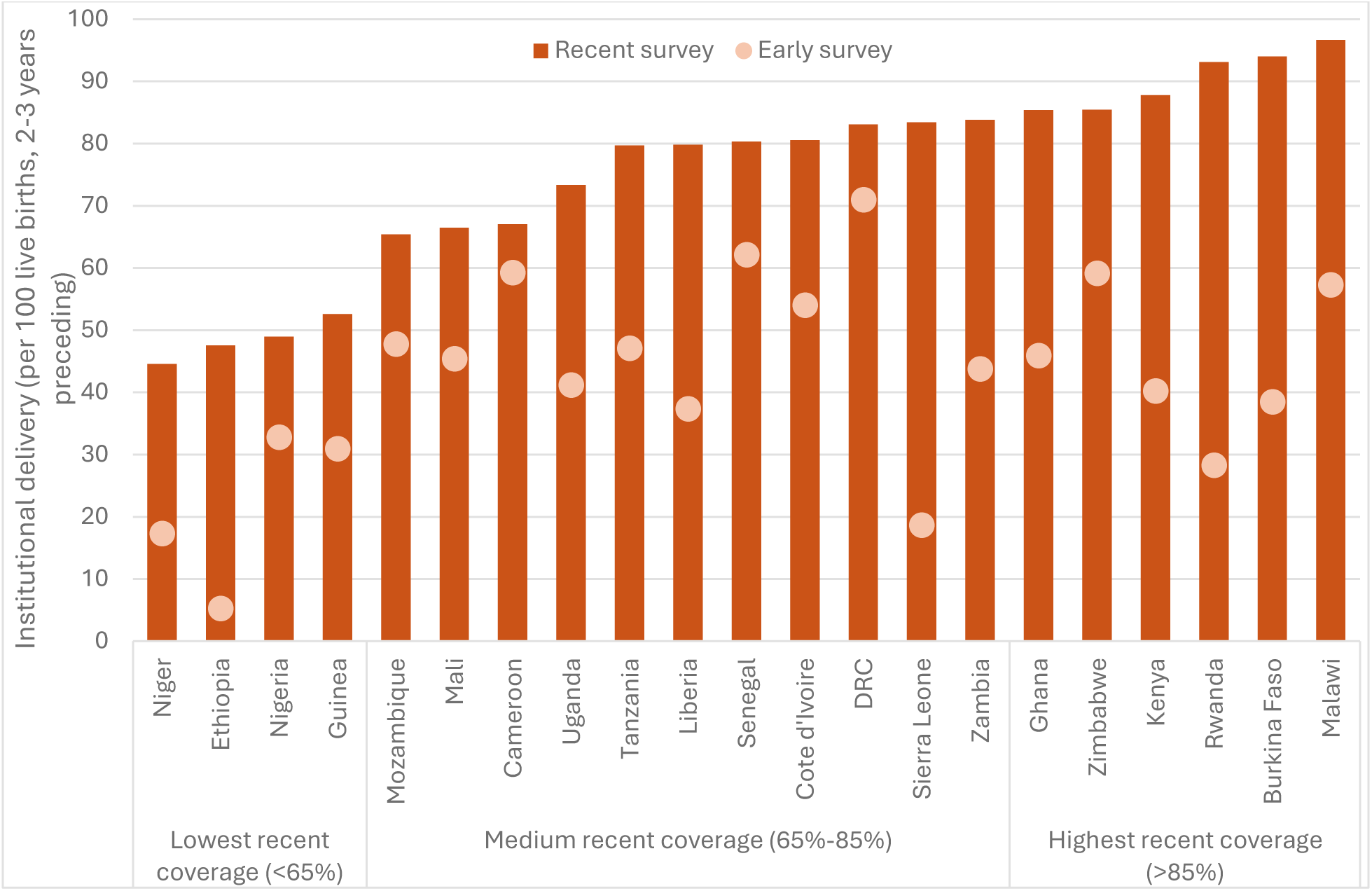
Trends in institutional delivery rates in 21 SSA countries (early and recent DHS/MICS, median years 2005-2019)

We next assessed trends in delivery coverage in more detail, including whether increases in institutional delivery rates in the lowest to highest coverage groups were increasingly marked by lower absolute rich-poor gaps, more births at hospitals (including among the poorest), and higher CS rates among the poorest (Table 1).

**Table 1:** Median coverage in early (median 2005) and recent (median 2019) surveys, by institutional delivery coverage group in recent surveys

| Coverage indicator | Countries with lowest coverage (<65%) | Countries with medium coverage (65-85%) | Countries with highest coverage (>85%) | All |
| --- | --- | --- | --- | --- |
| Institutional delivery (Phase IV median 96.9, IQR 87.7-98.6) |  |  |  |  |
| Early survey (median %) | 24.1 | 47.1 | 43.1 | 43.8 |
| Recent survey (median %) | 48.3 | 79.8 | 90.5 | 80.3 |
| Difference in medians (pp) | 24.1 | 32.7 | 47.4 | 36.5 |
| Institutional delivery, poorest (Phase IV median 91.8, IQR 69.0-96.9) |  |  |  |  |
| Early (median %) | 8.2 | 27.4 | 19.6 | 20.1 |
| Recent (median %) | 23.4 | 64.2 | 79.3 | 64.2 |
| Difference in medians | 15.1 | 32.7 | 59.8 | 44.1 |
| Institutional delivery, absolute rich-poor gap (Phase IV median 7.8, IQR 1.2-28.9) |  |  |  |  |
| Early (median pp gap) | 56.0 | 54.6 | 57.3 | 56.6 |
| Recent (median pp gap) | 64.1 | 31.3 | 18.4 | 31.3 |
| *Difference in median gaps (pp)* | 8.1 | -23.3 | -38.9 | -25.3 |
| Hospital delivery (Phase IV median 72.8, IQR 57.0-94.4) |  |  |  |  |
| Early (median %) | 11.5 | 24.0 | 30.3 | 24.0 |
| Recent (median %) | 13.7 | 29.5 | 51.1 | 29.5 |
| *Difference in medians (pp)* | 2.2 | 5.5 | 20.9 | 5.5 |
| Hospital delivery, poorest |  |  |  |  |
| Early (median %) | 2.1 | 6.6 | 11.7 | 6.6 |
| Recent (median %) | 2.5 | 12.7 | 32.1 | 12.7 |
| *Difference in medians (pp)* | 0.4 | 6.1 | 19.5 | 6.1 |
| C-section rate, poorest (population) (Phase IV median 15.1, IQR 7.4-21.9) |  |  |  |  |
| Early (median %) | 0.2 | 0.7 | 1.3 | 0.7 |
| Recent (median %) | 1.2 | 2.6 | 5.2 | 2.6 |
| *Difference in medians (pp)* | 1.0 | 1.9 | 3.9 | 1.9 |
| C-section rate, poorest (institutional) (NA in transition model) |  |  |  |  |
| Early (median %) | 2.9 | 3.5 | 5.6 | 4.0 |
| Recent (median %) | 5.4 | 3.9 | 7.3 | 4.8 |
| *Difference in medians (pp)* | 2.5 | 0.4 | 1.7 | 0.8 |

Improvements in institutional delivery rates were faster among countries that reached higher coverage levels. Positively, the overall absolute gap in institutional delivery coverage between the richest and poorest quintiles greatly declined by 25.3 points, from 56.6 pp to 31.3 pp from 2005 to 2019, respectively. However, when comparing coverage groups, this occurred among countries with the medium and highest coverage, but not yet in the lowest coverage group. Moreover, to reach Phase IV’s typical value of a 7.8 pp rich-poor gap will require ongoing improvements in all countries, especially among the poorest to reduce these gaps.

Increases in institutional delivery coverage were driven more by increases at LL facilities compared to hospitals, as hospitals still accounted for a smaller proportion of all deliveries (29.5%, versus 45.0% at LL facilities in recent surveys). These trends also diverged by coverage group, as the increases in hospital deliveries contributed more than 25% of the overall increase in five of the six higher coverage countries, in three of 10 medium coverage countries, and none of the four lower coverage countries. By the recent surveys, the highest coverage group’s median hospital delivery rate (51.1%) was considerably higher than those of the medium and lower coverage groups of countries (29.5% and 13.7%, respectively), though all were still below Phase IV’s median of 73%.

The increases in the proportion of LL facility births were more consistently high across countries among the poorest group than the population overall, or especially in the richest group (R-squared of 0.92 versus 0.75 and 0.28 respectively). Increases in hospital deliveries were lower overall and in the richest groups (R-squared of 0.40 each), and slightly more in the poorest groups for some countries (R-squared of 0.53) (Figure 2). Compared by recent coverage group, hospital deliveries in the poorest group improved most noticeably among countries that reached higher overall institutional deliveries (from 11.7% to 32.1%) (Table 1).

**Figure 2:**
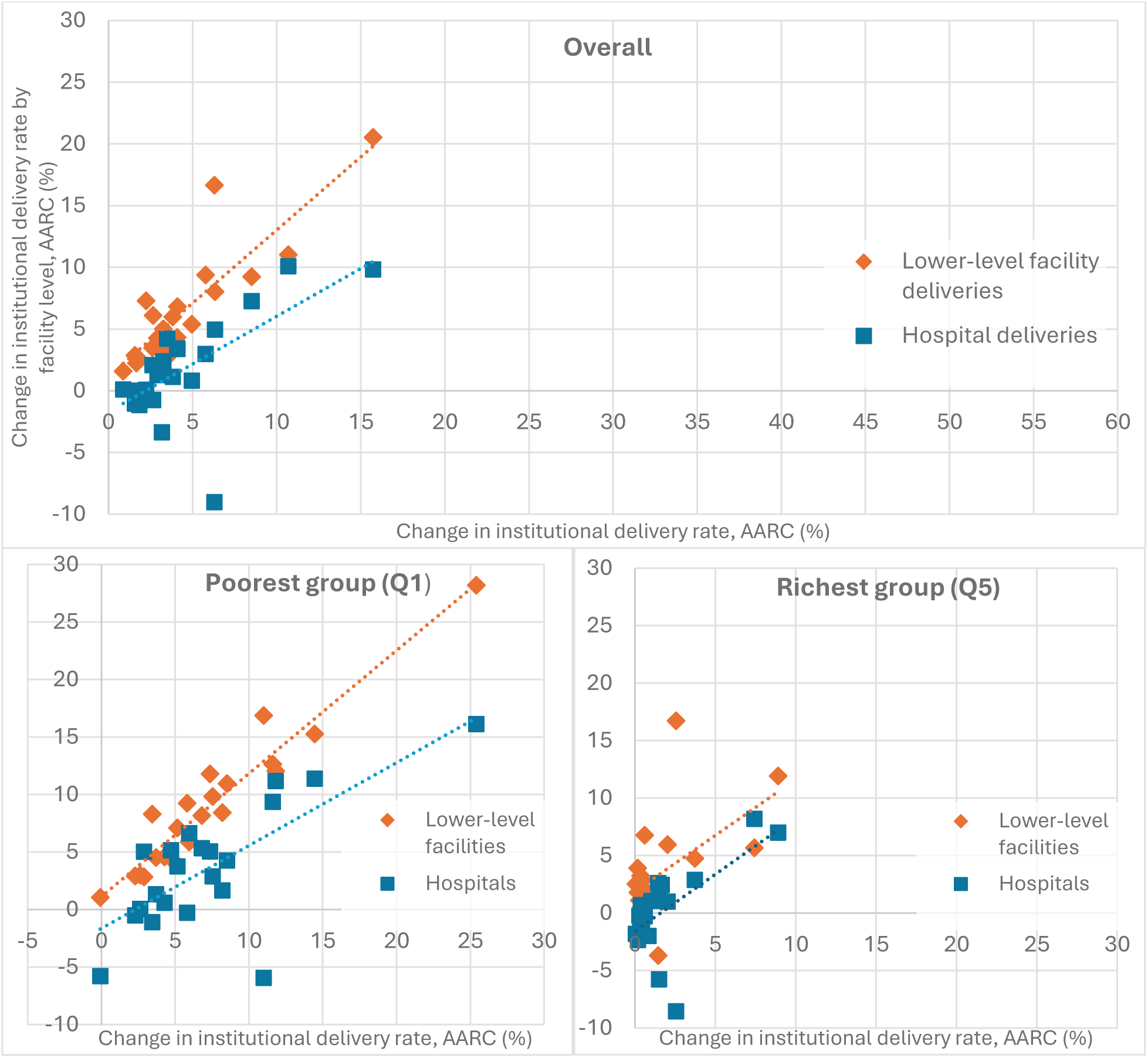
Countries’ average annual rates of change (%) in institutional deliveries at hospitals and LL facilities, overall and among the poorest and richest groups (from early to recent DHS/MICS)

Turning to CS, population rates started at 5% or below everywhere, and even lower among the poorest, which increased to varying degrees over time (Table C.1). It is more critical to examine CS rates among the poorest, who usually have very low rates of non-medically indicated procedures, as a better indicator of unmet need for CEmONC. Population CS rates among the poorest increased from a median of 0.7% to 2.6%, with the greatest increase in the highest coverage group (from 1.3% to 5.2%). This was still well below the 10% recommended by WHO, and 15.1%, the median for Phase IV countries (26). Institutional CS rates among the poorest increased little (from 4.0% to 4.8% for all countries). However, the median institutional CS rate increased most in the lowest coverage group, which was mainly due to large increases in Ethiopia (to 8.1%) and, to a lesser extent, Nigeria (to 6.0%), suggesting that more poor women with emergencies reached facilities capable of conducting CS.

We next examined whether more births were attended by skilled providers at each facility level, comparing changes between the three coverage groups of countries (Figures C.1 shows country-wise results). Among births at LL facilities, the greatest gains occurred in attendance by nurses or midwives in all coverage groups of countries but especially among the lowest coverage group (Figure 3). The mean proportion of births delivered at a LL facility by a doctor remained low in all groups. In contrast, the mean proportion of deliveries by nurses and midwives increased only slightly among hospital births. Birth attendance by a doctor increased most at hospitals, though this was only noticeable among the highest coverage group of countries (from a mean of 5% to 19% of all institutional deliveries). Births by other skilled providers at LL facilities still occurred but were a diminishing proportion of the overall increase in all groups.

**Figure 3:**
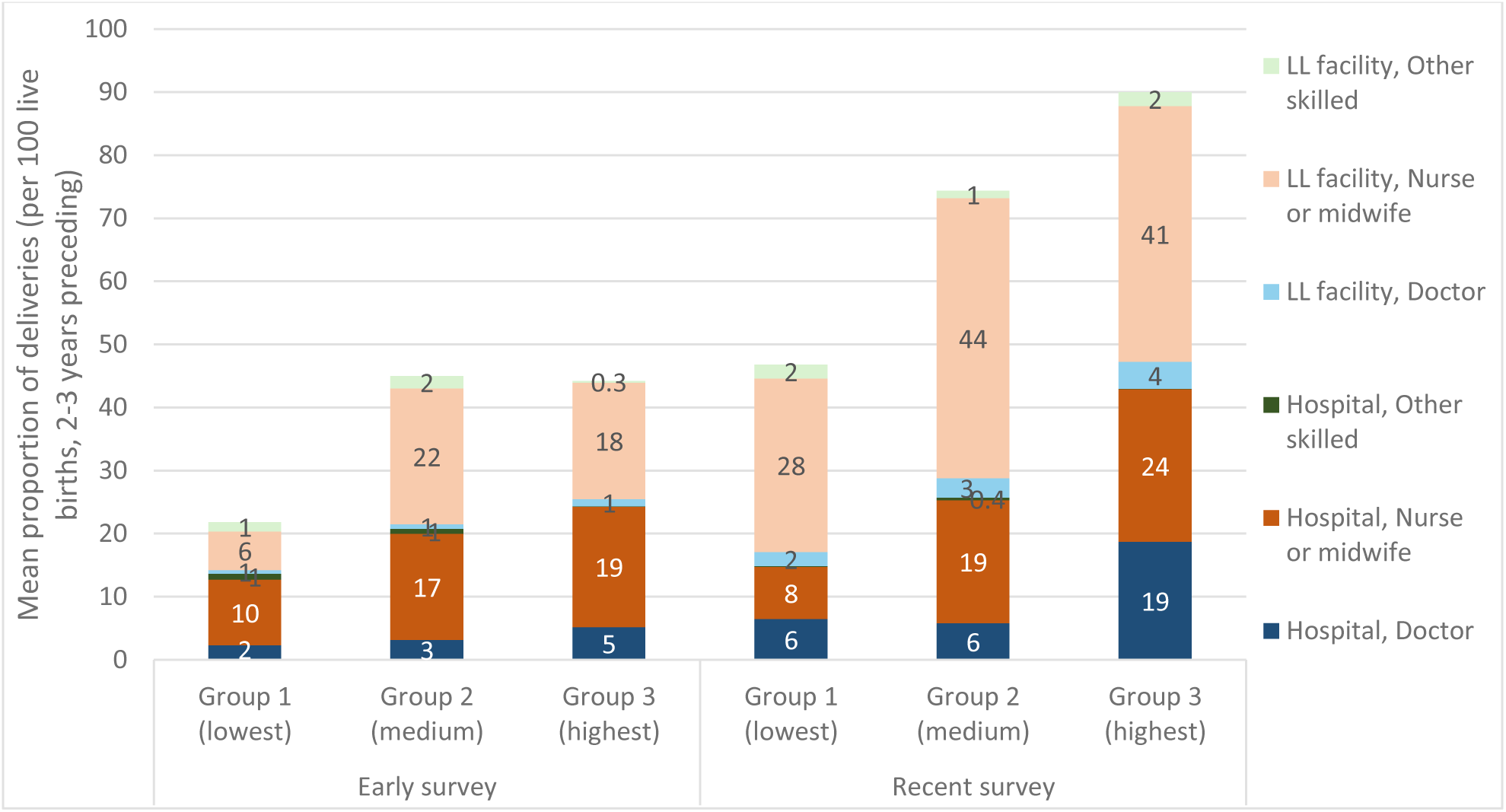
Mean proportion of deliveries by birth attendant type at hospitals and LL facilities, by country coverage groups (early and recent DHS/MICS)

We then examined skilled birth attendance (SBA) type at all locations by household wealth. Attendance by a doctor among the poorest women increased only in the highest coverage group of countries (from a mean of 3% in 2005 to 15% in 2019) (Figure 4, Figure C.2 shows country-wise results). In contrast, the mean proportion of the richest group attended by a doctor increased equally across coverage groups. On average, at least twice as many women in the richest group received delivery care from a doctor than the poorest group. Births attended by nurses or midwives among the poorest group increased most in all three coverage groups on average, to the level of the richest in the medium and highest coverage groups (over 60%). The mean proportion of the richest women attended by nurses or midwives remained high and increased only among the lowest coverage group of countries (from 47% to 62%). Attendance by other skilled providers only contributed to rising institutional delivery rates among the lowest coverage group (from a mean of 7% to 26%), but a declining and lower proportion among the medium and higher coverage groups of countries. For the richest, births by other skilled attendants declined from low levels, particularly among countries in the lower coverage group.

**Figure 4:**
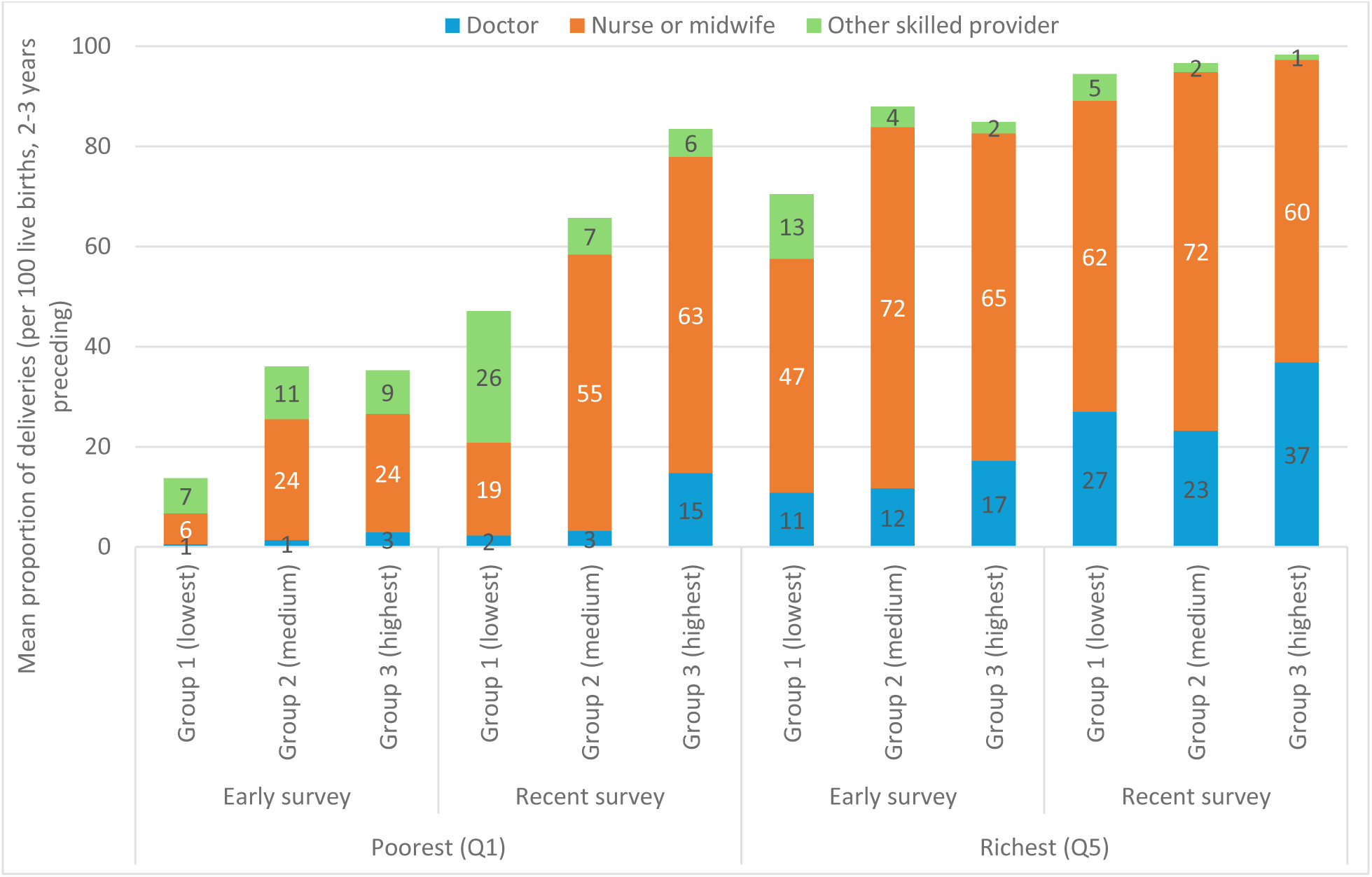
Mean proportion of deliveries by attendant type among the poorest (Q1) and richest (Q5) groups, by institutional delivery coverage group (early and recent DHS/MICS surveys)

### 3.2 What proportion of live births occurred at facilities with higher volumes of live births and CS in 2022 (HMIS/DHIS2)?

In most countries, the majority of annual live births took place in LL (or non-hospital) facilities in 2022. However, this pattern varied between coverage groups, as in the survey results. Countries in the lower and even medium recent coverage groups had medians of 20% of live births occurring at hospitals (except Zambia at 43%) and 80% at non-hospitals. For the highest coverage group of countries, the distribution was much more even with 52% of live births occurring at hospitals (except Burkina Faso at 15%) versus 48% at non-hospitals. Median live birth volumes per facility were generally higher in hospitals than non-hospitals (869 versus 114 respectively), but this varied greatly with inconsistent patterns across coverage groups (Figure 5a).

**Figure 5:**
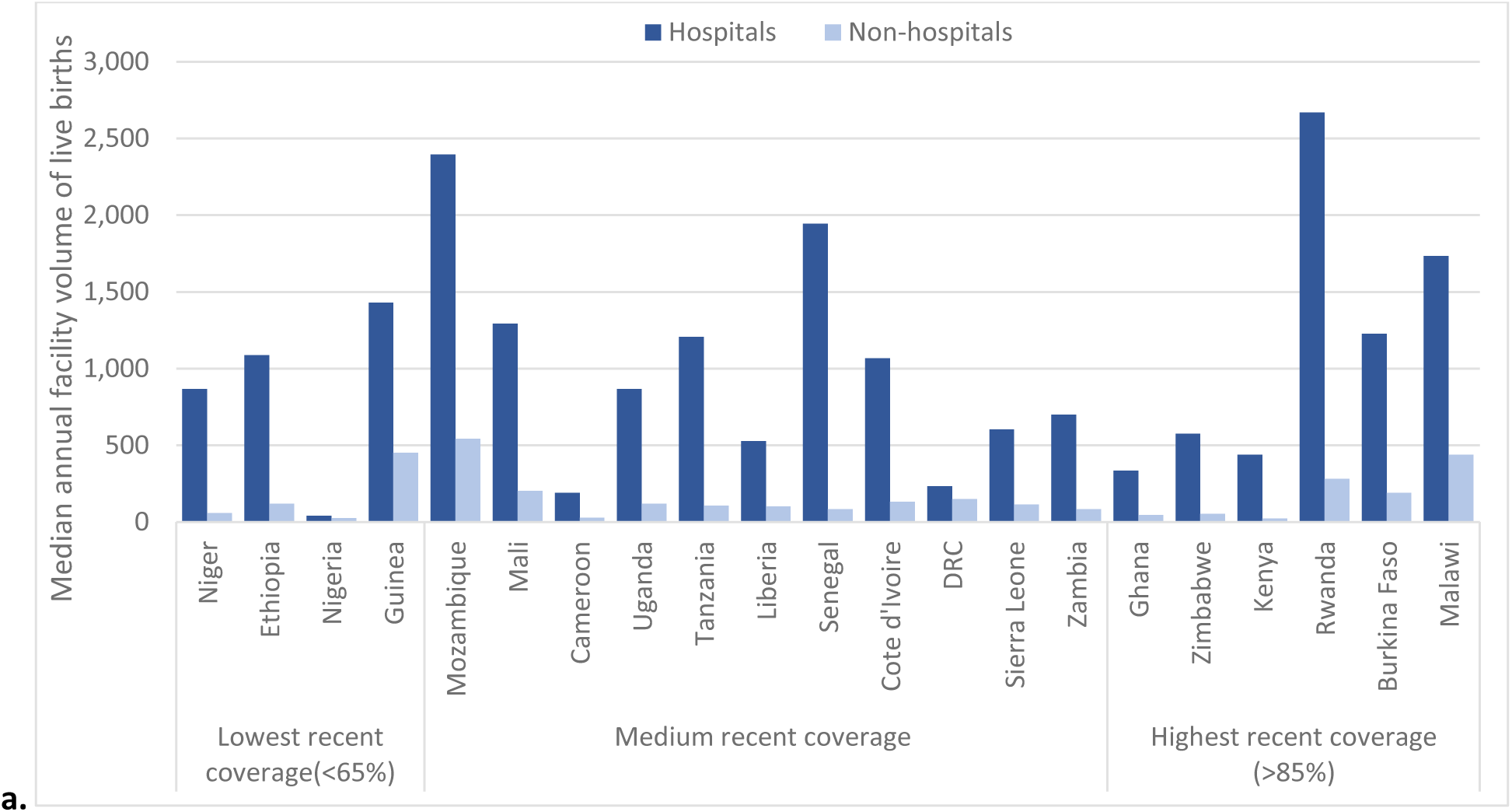
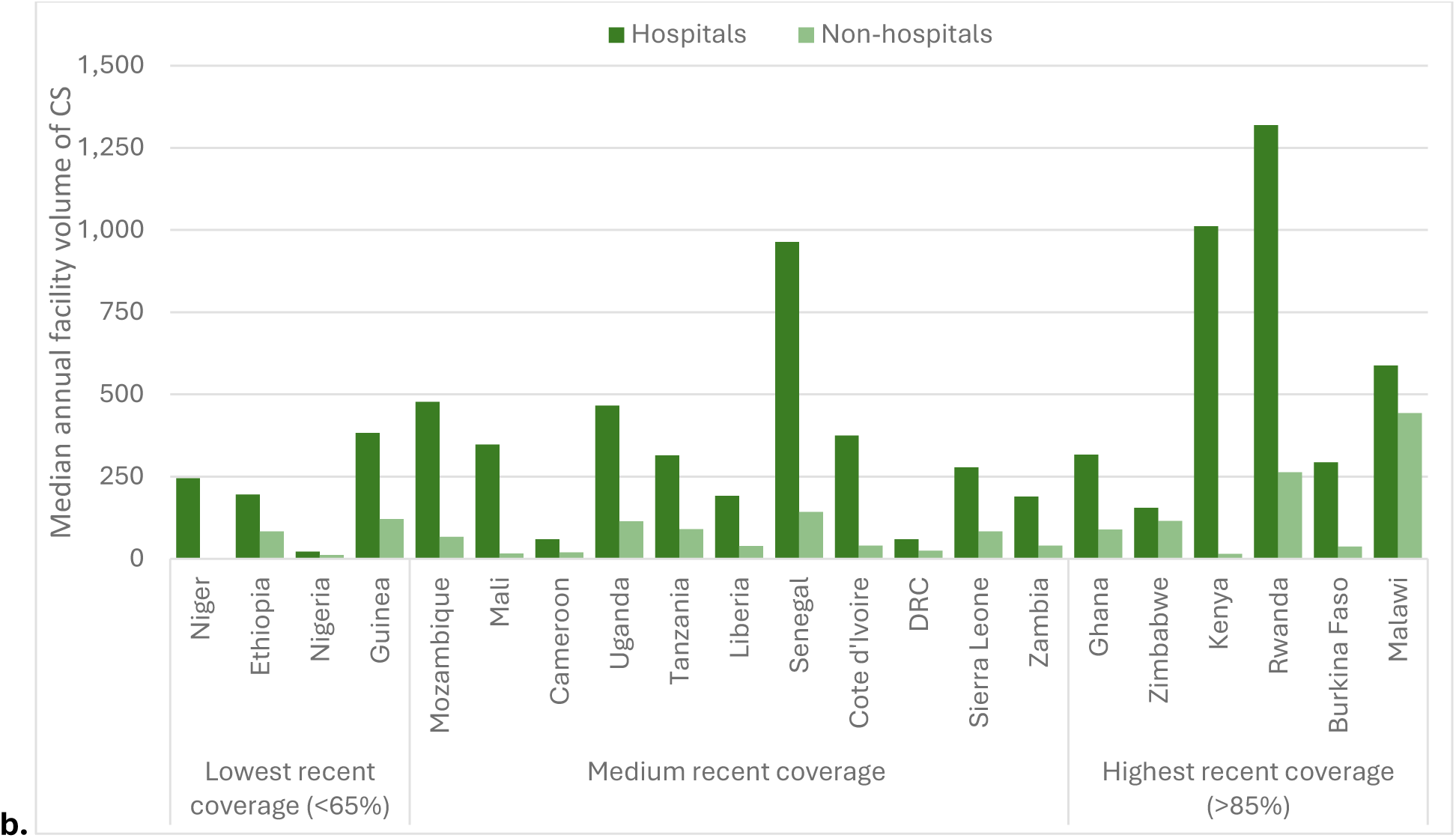
Median annual facility volume of (a) live births and (b) CS at hospitals and non-hospitals, by coverage group in the recent survey (HMIS 2022)

Unlike for live births, the proportion of CS at hospitals was consistently higher than at LL facilities across all coverage groups. The overall median proportion of facilities reporting any CS was 85% at hospitals and 2% at LL facilities. The median proportion of CS in hospitals among the lowest and medium coverage groups was about 80%, while for the highest coverage group nearly all CS (95%) occurred at hospitals. Only in some countries was the proportion of CS reported at non-hospitals slightly more noticeable (7-14% of CS), including Malawi, Tanzania, Cameroon, Uganda, Nigeria, Sierra Leone, and DRC (Figure C.3). There was wide variation in countries’ median annual CS volume per facility, among those with any CS (Figure 5b). This was consistently higher for hospitals than non-hospitals (315 versus 84 respectively), typically ranging from 200 to over 1000 per hospital annually. We excluded Niger data on lower-level facilities as there were very few facilities reporting any C-sections (0.2%) and a few facilities with high annual volumes distorted the picture for lower-level facilities, presumably due to misclassification (Table A.4).

Next, we examined the proportion of live births at facilities with lower and higher annual median volumes in 2022 by country and coverage group (Figure 6; Figure C.4). The proportion of live births reported at lower-volume non-hospitals (less than one every other day, or 183 per year) did not vary substantially across coverage groups. However, the median was slightly lower among countries with the highest delivery coverage compared to others (13%, compared to 17-19%, respectively). Conversely, the proportion at higher-volume hospitals (over one birth per day, or 500 per year) was much higher in the highest coverage group than medium and lower (median 49% versus 20% and 19% respectively). Although, this pattern was less evident in Burkina Faso, where the majority of births (55%) occurred at higher-volume non-hospitals with one or more live births per day, as well as in Sierra Leone, Cameroon, Nigeria, and Niger where 30% to 50% of births occurred at lower volume LL facilities.

**Figure 6:**
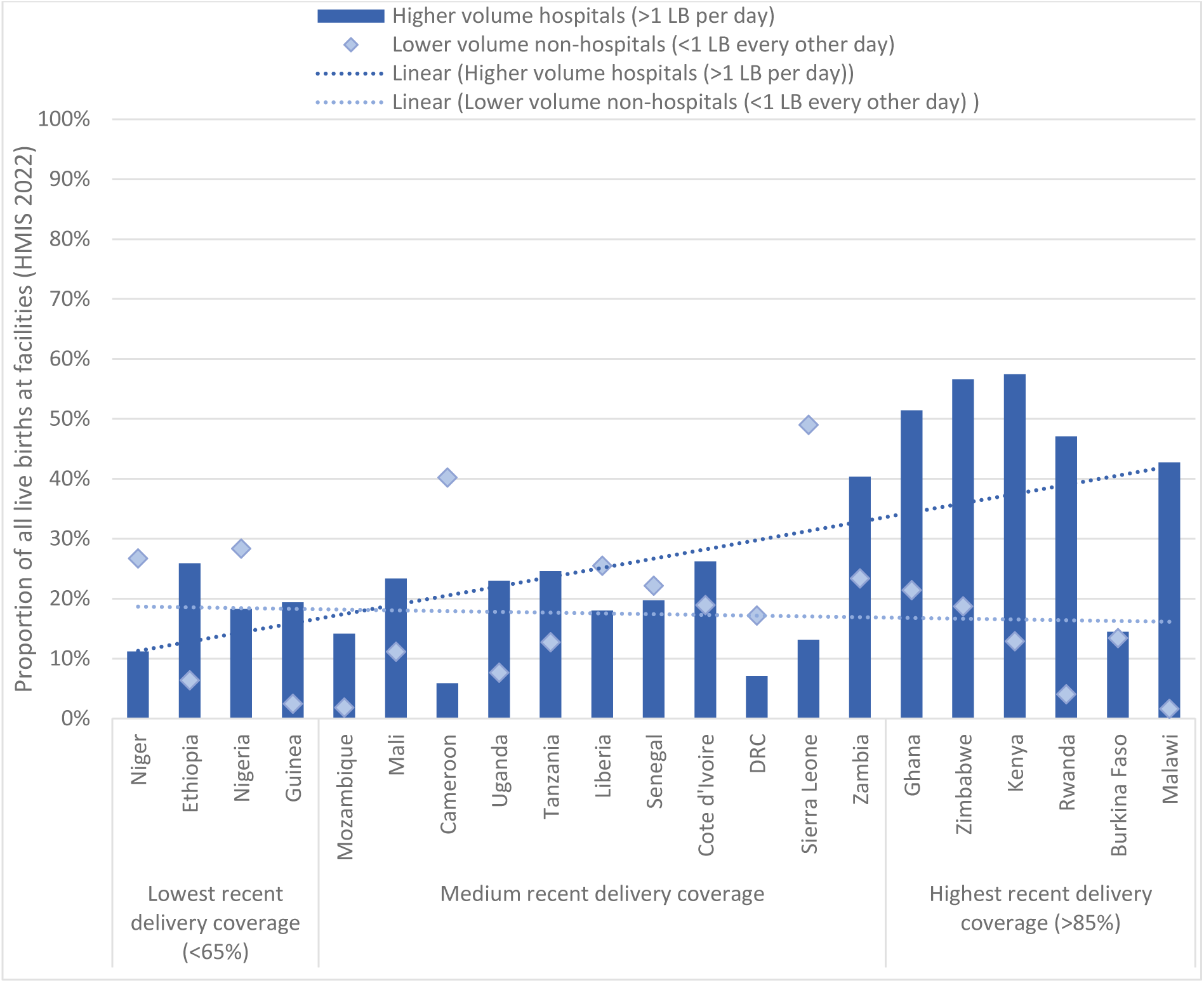
Proportion of all live births at lower versus higher median volume hospitals and non-hospitals (HMIS 2022), by institutional delivery coverage

**Figure 7:**
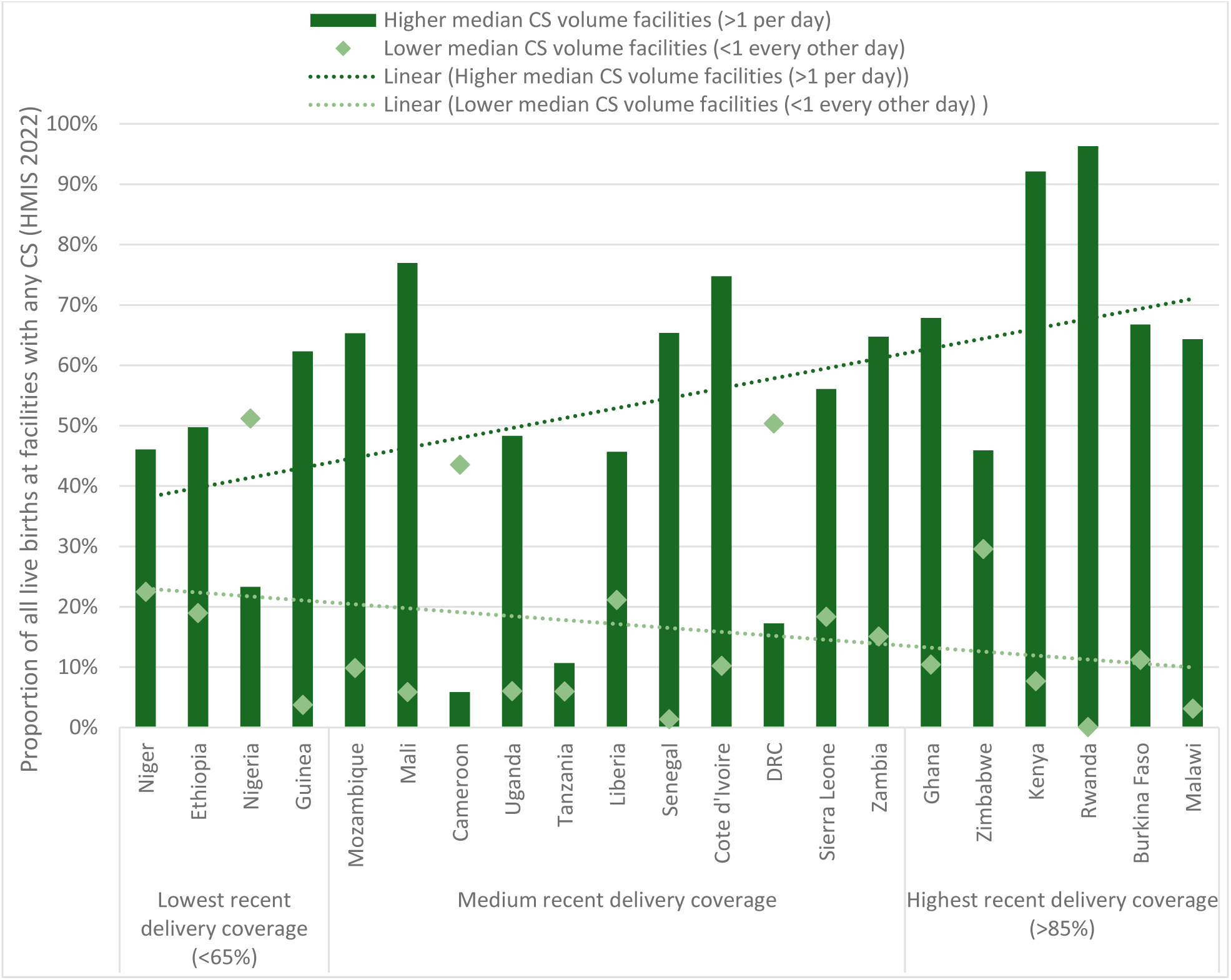
Proportion of live births at facilities performing any CS with lower versus higher median annual volumes of CS (HMIS 2022), by institutional delivery coverage

We examined the proportion of live births at facilities that conducted any CS with lower versus higher median annual CS volumes, as a proxy for the comprehensiveness and quality of childbirth care. We combined all facilities with any CS, regardless of level, given the relatively low proportion of CS at non-hospitals (Figure C.5 presents stratified results by non-hospital and hospital). We found that the proportion of live births at facilities performing a lower CS volume (less than one every other day, or 183 in a year) was generally low except in a few countries (Nigeria, Cameroon, and DRC). The proportion of live births at lower CS volume facilities were somewhat lower as countries’ overall coverage levels increased (10% of births in the highest compared to 21% in the lowest coverage group). Conversely, the proportion of live births at higher CS volume facilities (more than one per day, or 500 per year) became somewhat higher among higher compared to lower delivery coverage group of countries (median of 67% versus 48%, respectively).

## 4. Discussion

This study sought to better understand trends in facility-based childbirth care over the past two decades in 21 SSA countries. To date, the rapid rise in institutional delivery rates has only partially translated into equitable access to comprehensive, life-saving childbirth care provision in 21 countries of SSA. Countries that reached the highest recent coverage experienced the greatest gains overall and among the poorest women. Consistent with the MNH transition model, the proportion of births occurring at hospitals, with more skilled attendants and higher volumes, and to some extent greater CS capacity, generally rose across countries with increasing institutional delivery coverage as well. We now consider the findings in light of other evidence, study countries’ policies, health system inputs and recommendations, and wider debates to guide future MNH care strategies to ensure equitable access and quality of childbirth care in SSA.

The rapid rise in institutional delivery rates across SSA kept pace with the large absolute growth in the number of births (17), with the greatest increases continuing to be at LL facilities that each conducted a lower volume of births across countries (3). Past studies showed that inequalities in institutional delivery rates have reduced in most SSA countries, but that subnational and socio-economic disparities still exist particularly for hospital delivery rates (17,27). Yet the observed trend towards more hospital births overall and among the poorest in countries reaching higher institutional delivery coverage is encouraging, given that LL facilities are less able to handle higher volumes or emergencies, often with insufficient referral linkage to CEmONC facilities (15). Moreover, some studies showed that the quality of childbirth care at non-hospitals (even with higher birth volumes) was lower than at hospitals, and especially hospitals with CS capacity (7,8,12).

Skilled birth attendance also reached higher levels, with a now negligible role of skilled attendants at home, compared to past evidence (28). To date, SSA generally lagged behind other regions, with significant intra-country inequities due to a range of factors (15,17,27–29). Higher transition phases are typically characterized by greater health worker densities, with doctors relatively increasing compared to nurses or midwives. Our findings confirm that birth attendance by nurses or midwives – but not doctors – increased at LL facilities, while the proportion attended by doctors at hospitals rose to nearly the same as nurses and midwives. This is significant given that doctors and medical officers are the primary cadres trained to perform CS in most study countries. The proportion of the poorest women attended by doctors also only increased for countries that reached higher institutional delivery coverage. Attendance by doctors was consistently higher among the richest, even in the countries with lower coverage levels, reflecting ongoing inequities by SBA type (16,30).

C-section rates in the 21 SSA countries remained below five percent among the poorest, and even in the general population in some countries, which is lower than global averages (18,31). Only the highest coverage group saw noticeable increases among the poorest, but rates remained below 10% as recommended by WHO for met need, except in Rwanda and Ghana (10,16,19,31–33). Prior research has found that wealthier women had higher rates not only of elective but also emergency CS, underscoring that poorer women are less able to access CS when needed (18,34). Indeed, SSA now faces a dual challenge of underuse of CS in rural and poorer populations and potential overuse in urban and wealthier populations (18,32,34–37). Little improvement in institutional CS rates among the poorest women across countries suggests that health facilities could not keep up with the rising need for life-saving emergency care as more of them started delivering at facilities (33). Some non-hospitals conducted CS, but most with very low volumes. While LL facilities may fill a gap for CEmONC where hospitals are scarce, maldistributed, or underutilized, their low CS volumes also challenge its quality. Across countries, most CS occurred at hospitals that had high birth volumes, but this generally constituted a greater proportion of live births among countries with higher than lower overall delivery coverage as well.

In SSA, studies have found that facility choices for childbirth have been driven by a complex combination of maternal age and birth order, education, wealth, socio-cultural and gender norms, distance and travel time, health system constraints, environmental barriers, conflict, and natural disasters (15,27–29,35,37–42). The sharp rise in births at LL facilities may have stemmed from the explicit dual policy on childbirth service organization to direct births assessed to be lower risk to LL facilities and those assessed as higher risk to hospitals in most of the study countries (38). This has been found to be a more cost-efficient approach, but not the most effective in averting poor outcomes (43), given the challenges of accurately assessing risk during pregnancy and transporting emergencies to higher levels without delays (11,15,18). In this study, many countries’ policies focused on increasing community demand for SBA at facilities and establishing more LL health facilities especially in rural areas. This has lowered distances and travel times for LL facilities (38). A study in four SSA countries estimated an average of over one-hour of travel time to hospitals, above the 30 minutes now recommended by WHO in cases of emergency referrals (35). There is also more maldistribution of hospital densities compared to LL facilities at the subnational level (39). In countries’ 2022 HMIS data, many neared the global target of two health facilities per 10,000 population overall (with a median of 1.7), with the majority being LL facilities. Countries with the highest coverage had a slightly higher median density of hospitals than countries with lower overall coverage. However, the densities of all facilities and human resources were still lower in most remote or rural districts everywhere, alongside growing gaps in highly populated urban areas.

Health policy and system reasons for low and inequitable CS rates may also include insufficient infrastructure, a lack of trained health workers, and inadequate required equipment, all exacerbated by socio-economic barriers (16,32,34). Study countries indicated that the distribution of EmONC was particularly insufficient and inequitable, while health worker capacity for managing complications was widely lacking especially at LL facilities. Women and families sometimes voluntarily bypass LL facilities due to higher perceived readiness or EmONC functionality at hospitals, also reflecting maldistribution of facilities with sufficient capability and quality (16,35). Other studies in a few SSA countries found that despite having CS capacity, some hospitals had insufficient EmONC signal functions, which deserves further attention (29,44,45). Many study countries’ essential medicine or commodity lists include key MNH products, and some have put central distribution systems in place. Some have also recently adopted policies on EmONC referral networks and emergency transport. Yet they indicated that implementation has often been constrained by uncoordinated communication systems, insufficient public ambulances, limited road infrastructure especially remotely, and inconsistent funding to support this.

Going forward, policy-makers, researchers and partners must refine their strategies to be increasingly multifaceted and contextualized to ensure high-quality MNH care that reduces mortality levels for all women and newborns in SSA (4,22). Optimizing the ‘division of labour’ for childbirth care requires continually understanding the country’s evolving MNH landscape. This means actively tracking delivery coverage patterns and birth volumes by facility type vis-à-vis the population in need at more granular levels – disaggregated by wealth, geography, and health service functionality – to improve the densities and distributions of equipped health infrastructure (3,4,16,34,38). Investments must be guided by real-time data, inclusive planning processes, and accountability mechanisms to ensure responsiveness to local needs and sustainability. The need for such contextualization aligns with global frameworks such as networks of care and system redesign, which emphasize ensuring continuity, quality, and integration of MNH care, enhancing linkages, combined with strategically upgrading facilities to provide more comprehensive childbirth care (11,12,46).

While there had been debates on whether to phase MNH policies, starting with availability and demand and then improving quality, this study’s findings underscore the need to redouble efforts to prioritize both effectiveness and equity in access to life-saving MNH care at once (4,12,15,16). Neglecting this would risk entrenching inequities in the health system, particularly if families perceive care as unsafe, disrespectful, or ineffective. Study countries at all coverage levels still recommended continuing community education and outreach to promote demand for facility-based delivery care, while increasing availability through task-shifting and incentives for rural recruitment and retention, focusing on disadvantaged populations where coverage remains lower (47). Additionally, most countries have already established policies on quality of childbirth care in the last five or ten years, including some on experiences of care like in Ethiopia. Countries that reached higher overall coverage also specifically recommended quality improvements in facilities serving more disadvantaged groups, through more consistent supports, supervision and in-service training for providers around basic emergency obstetric and neonatal care (BEmONC), newborn resuscitation, and respectful maternity care.

Improving access to high-quality CEmONC requires even more intensive resources that can strain a health system (7,8,44,48,49). Yet this seems crucial to move towards Phase IV with more equitable CS rates (4,18,34,48). Some countries across coverage levels have started expanding CEmONC facilities such as Ethiopia, Côte d’Ivoire, Mozambique, and Zambia, and newborn care units in more districts in Guinea and Mali. Another strategy is upgrading LL facilities to offer CEmONC (38,50). Half of the study countries recommended this approach, and Tanzania has started; the need to evaluate its cost-effectiveness was also noted. Some argue that concentrating available resources in fewer, but strategically selected, facilities may thus promote accessibility for all (38). To facilitate access, countries indicated that their recent policies and guidelines on emergency referral transport and networks must be implemented with sufficient funding and coordination, consistent access to ambulances while minimizing distances and travel time, especially for remote areas. Other strategies include residence support before and after delivery such as maternity waiting homes or spaces for families to stay at hospitals (4,47).

Moving towards transition Phase IV has typically also been underpinned by more health financing and user fee removals (4). In terms of financing, the median total health expenditure in 2022 among study countries with lowest to highest coverage is estimated to have increased from US $41 to $74 per capita respectively (51). The former is comparable with typical median levels of Phase II countries, while the latter with Phase III levels, though typical Phase IV levels are substantially higher at over US $300 per capita (22). A greater proportion of these funds must be dedicated to MNH programming, which currently varies greatly among study countries between public, private or external sources (4,22,47,51). In terms of reducing user fees, the proportion of the total health expenditure that was out of pocket is estimated to be 50% to 18% among the lower to higher coverage countries respectively (51). In fact, the latter fell in the range of typical Phase V levels. It is laudable that nearly all 21 countries adopted policies to remove user fees in public facilities for vaginal as well as CS births in the past decade or more. Despite these policies, they emphasized that low CS rates especially for the poorest may partly relate to indirect costs, and in some countries, a growing reliance on private care due to higher perceived quality (43,47,52). Targeted subsidies or travel vouchers may help in the short term, but more sustainable financing mechanisms alongside system strengthening are needed for long term impact (30,31).

In a nutshell, sustained improvements in maternal and newborn survival in SSA will require full commitment to a paradigm shift – positioning equity and quality as non-negotiable pillars of investment across all phases of transition. Achieving this requires institutionalized accountability mechanisms, integrated subnational and national RMNCAH information systems, resilient financing, and multisectoral alignment with broader development priorities (4,53). Numerous study countries recommended strengthening monitoring, learning, and evaluation processes to use routine data and establish dashboards to track progress in real time, as well as conduct in-depth analyses of policy implementation challenges, to refine their strategies (4,47,54). Future efforts must prioritize context-sensitive, data-driven planning and evaluation frameworks. This includes pathway analyses to unpack persistent barriers to equitable access and quality, even in settings with higher coverage, while empowering local leadership and communities to shape and sustain high-quality, inclusive MNH care systems.

This study had some limitations. Definitions of facility and SBA types differed across year, data source, and country; we used countries’ definitions and policies to refine them. Wealth indices may have differed by country and over time, although it allowed comparisons of inequalities not possible with more complex measures. There may have been some recall bias particularly for birth attendant type in the surveys. Coverage and volumes were measured among live births rather than all births, due to underreporting of stillbirths. The use of arbitrary cut-offs for higher or lower annual birth volumes may be misleading, as the thresholds do not necessarily reflect variations in facility capacity or differences in population size across the different countries. However, when we compared proportions of births at facilities above or below an annual median volume of 500 to 1500, very similar patterns for countries were visible that led to similar conclusions. We also did not consider the timing of CS as before or after onset of labour; this may not be directly indicative of a medically-indicated CS and the rates of both appeared low elsewhere (18). Optimal population CS use is not universally agreed, but the WHO now says 10% is the maximum for additional health benefits, previously this was up to 15% (26). It was beyond the study’s scope to assess EmONC readiness, or to compare different hospital levels, but some countries have planned follow-up analyses.

## 5. Conclusion

This study aimed to better understand the major increases in institutional delivery over the past two decades in 21 SSA countries and the implications for health system strategies going forward. The rise in countries’ coverage levels was driven by increases in deliveries at LL facilities attended by nurses and midwives, followed by an increasing share at hospitals with doctors which had higher volumes and CS capacity. Lingering inequities exist in access to hospitals and CEmONC in most countries, despite improvements in those reaching higher coverage levels. These changes in delivery care patterns are largely consistent with the maternal, stillbirth and neonatal mortality transition model, which is based on data from 151 countries during 2000-2020. For countries to transition towards lower mortality levels requires optimizing the density, distribution, quality and linkages for comprehensive childbirth care to meet their evolving population needs at subnational levels. Countries have established a wide set of policies, leaving them poised to pursue effective and equitable progress, yet operational and resource challenges remain. Policy-makers, researchers, and implementers must co-develop and track increasingly multi-faceted, contextualized implementation strategies that ensure equitable, high-quality MNH care where and for whom it is most needed. In doing so, countries can more effectively close persistent childbirth care gaps that undermine progress towards leaving no women or newborn behind in SSA.

## Data Availability

The publicly available survey data for this article are available at https://mics.unicef.org/surveys and https://dhsprogram.com/Data/. The data from HMIS/DHIS2 was obtained by country collaborators that participated in the study. Those interested in replicating the analysis must contact countries for permission to access the datasets, upon reasonable request, by requesting the corresponding author to provide connection to the appropriate contact.

## Acknowledgements

We acknowledge the Countdown to 2030 collaboration and African Population Health Research Centre Administrative and Communications teams for the invaluable support to this multi-country study.

