## Supplemental materials for "Transitions in childbirth care provision: Understanding the rapid rise in institutional delivery in 21 countries of sub-Saharan Africa and the implications for future strategies"

### Supplementary materials

#### Appendix A

##### Table A.1: Survey type and year analyzed in the study*

| Country | Early survey type and year | Recent survey type and year |
|---|---|---|
| Burkina Faso | DHS, 2003 | DHS 2021 |
| Cameroon | DHS 2004 | DHS 2018 |
| Cote d'Ivoire | MICS 2006 (2011 for C-sections)** | DHS 2021 |
| DRC | DHS 2007 | MICS 2017 |
| Ethiopia | DHS 2005 | DHS 2019 |
| Ghana | DHS 2003 | DHS 2022 |
| Guinea | DHS 2005 | DHS 2018 |
| Kenya | DHS 2003 | DHS 2022 |
| Liberia | DHS 2007 | DHS 2019 |
| Malawi | DHS 2004 | MICS 2019 |
| Mali | MICS 2006 | DHS 2018 |
| Mozambique | DHS 2003 | DHS 2022 |
| Niger | DHS 2006 | DHS 2021 |
| Nigeria | DHS 2003 | MICS 2021 |
| Rwanda | DHS 2005 | DHS 2019 |
| Senegal | DHS 2005 | DHS 2019 |
| Sierra Leone | MICS 2005 (2008 for C-sections)** | DHS 2019 |
| Tanzania | DHS 2004 | DHS 2022 |
| Uganda | DHS 2006 | DHS 2016 |
| Zambia | DHS 2001 | DHS 2018 |
| Zimbabwe | DHS 2005 | MICS 2019 |

*****The surveys used standardized questionnaires to collect information on various health topics, as well as individual, household, and community characteristics. These surveys are usually conducted by a designated national statistical agency. Ethical approval was obtained from the institutions responsible for commissioning, funding, or conducting the surveys. **C-section data was not available in prior survey used for the other analyses, so the next survey was used for those analyses.

##### Table A.2: Survey facility and birth attendant type categorizations

| Country | Survey type & year | Place of delivery category | | | Birth attendant | | | |
|---|---|---|---|---|---|---|---|---|
| | | Hospital | Lower level facility | Home or other | Doctors | Nurse/ Midwife | Other skilled provider | Unskilled provider/ none |
| Burkina Faso | DHS 2003 | Hôpital, Hôpital/clinique privé | Centre de santé, Autre privé médical (préciser), Autre public (préciser) | Votre domicile, Autre domicile, Autre | Médecin | Infirmiére/ Sage-femme/ Assistant medical | Accouchese traditional formée | Accouchese traditional non formee, Parents/Amis, Autre (precisér), Personne |
| | DHS 2021 | hôpital gouvernemental, Hôpital/clinique privé | Maternité, centre de santé/ PMI, Autre secteur med. Privé | Votre maison, Autre maison, Autre | Médecin | Infirmiére/ Sage-femme | Accoucheuse Auxiliaire, Matrone/ Accouc. Formée | Accoucheuse Traditionelle, Agent de Santé Communaut/ Village, Guérriseur traditionnel, Ami/Parents, Autre, Personne |
| Cameroon | DHS 2004 | Hôpital, Hôpital privé confess, | Centre de santé, Centre Santé/Dispens./C | Votre domicile, Autre | Médecin | Infirmér(e)/ sage-femme | Sage-femme auxiliaire, | Accoucheuse traditionnelle, |

| | | | | | | | | |
|---|---|---|---|---|---|---|---|---|
| | | Hôpital privé laïc/Clinique | onfess./Mission, Autre privé médical (Préciser) | domicile, Autre | | | Aide Soignante | Amis/Parents, Autre, Personne |
| | DHS 2018 | Hôpital public, Hôpital/ Clinique confessionel, Hôpital/ Clinique privé Laic | Centre de santé intégré/ dispensarie public, Centre médical D'arrondissement, Autre secteur public, Centre de santé /dispensaire/confessionel, Cabinet médical, Autre secteur médical privé | Sa maison, Autre maison, Autre | Médecin | Infirmére / Sage-femme | Aide-soignant | Accoucheuse traditionnelle, Amis/Parents, Autre, Personne |
| Côte d'Ivoire | MICS 2006 | Hôpital public, Hôpital privé, Clinique privé | Clinique/ centre de santé public, Autre public, Maternité privé, Autre médical privé (préciser) | Votre domicile, Autre domicile, Autre | Medécin | Infirmier(e)/Sage-femme | Aide-soignante/Sage-femme auxiliaire, Accoucheuse traditionnelle, Agent de santé communautaire, Autre: Tavailleurs sociaux | Parente/Amie, Personne |
| | DHS 2021 | CHU/Centre spécialisé Gouv, CHR/Hôpital général, Hôpital privée | Centre de santé Gouv. (ESPC), Autre public (préciser), Clinique / infirmerie privée, Maternité privée, Autre privé privé médical (préciser) | Domicile de l'enquêtée, Autre domicile, Autre | Médecin | Infirmière /Sage-femme | Aide-soignante, Accoucheuse traditionnelle, Agent de sante communautaire, Fille de salle, | Parente/Amie, Personne |
| DRC | DHS 2007 | Hôpital, Hôpital/ clinique privé | Centre de Santé, Poste de santé, Autre secteur public, Centre de santé privé, Autre secteur médical privé | Votre domicile, Autre domicile, Autre | Médecin | Infirmière, Sage-femme, Accoucheuse | Matrone de village, Accoucheuse traditionale | Guerriseur traditional, Autre, Maman du quart./Village, Personne |
| | MICS 2017 | Hôpital gouvernemental, Clinique / Centre de santé Gouv, Hôpital privé, Clinique privé | Poste de santé Governmental, Autre public (Préciser), Maternité privé, Autre privé médical (precisér) | Domicile de l'enquêtée, Autre domicile, Autre | Médecin | Infirmièr/ infirmiere, Accoucheuse/Sage-femme | Accoucheuse traditionnelle /matrone | Agent de santé communautaire, Parent(e) / Ami(e) , Autre (préciser), Personne |
| Ethiopia | DHS 2005 | Government hospital/Clinic, Private hospital/Clinic | Government health center, Government health post, NGO Health facility | Your home, Other home, Other | (Covered by Health professional) | (Covered by Health professional) | Health professional, Trained traditional birth attendant | Community health agent, Untrained traditional birth attendant, Other, No one |
| | DHS 2019 | Government hospital, Private hospital | Government health center, Government health post, NGO: health facility, Other public | Her home, Other home, Other | Doctor | Nurse, Midwife | Health officer, Traditional birth attendant | Health extension worker Other, No one |

| | | | | | | | | |
|---|---|---|---|---|---|---|---|---|
| | | | sector, Private clinic | | | | | |
| Ghana | DHS 2003 | Govt. Hospital/Clinic, Private hospital/clinic | Govt. Health Center, Govt. Health Post, Maternity home | Your home, Other home, TBA's home, Other | Doctor | Nurse/Midwife | Auxiliary midwife | Traditional Birth Attendant, Relative/Friend, Other, No one |
| | DHS 2022 | Government hospital, Private hospital | Government clinic/health centre, Government health post, Other public (specify), Private clinic, Private maternity home, Other private medical (specify), | Respondent's home, Other home, Other | Doctor | Nurse/Midwife | Community health officer/Nurse | Traditional Birth Attendant,Villager Health volunteer, Traditional Health Practitioner, Relative/Friend, Other, No one |
| Guinea | DHS 2005 | Hôpital, Hôpital/Clinique privé | Centre de Santé, Poste de santé, Autre privé médical | Votre domicile, Autre domicile, Autre | Médecin | Infirmère /sage-femme, Aide de Santé | Agent Technique de Sante (ATS), Accoucheuse formée, | Accoucheuse Traditionelle, Parents/AMIS, Autre, Personne |
| | DHS 2018 | Hôpital Gouvernement, Hôpital Regional, Hôpital Prefecture/ Centre Médical Communal (CMC), Hôpital/ clinique privée | Centre Santé Gouvt., Poste de Santé Gouvt., Autre secteur public, Clinique PF/AGBEF, Cabinet privé de Sage-femme | Sa maison, Autre maison, Autre | Médecin | Infirmiére/Sage-femme | Agent Technique de Sante (ATS) | Accoucheuse traditionelle, Agent de santé communautaire /Village, Parents/amis/voisins, Autre, Personne |
| Kenya | DHS 2003 | Govt. hospital, Private hosp/clinic, Mission hospital/clinic | Govt. health center, Govt. dispensary, Other public, Nursing/maternity home, Other private medical | Your home, Other home | Doctor | Nurse/Midwife | | Traditional birth attendant, Relative, friend, other, no one |
| | DHS 2022 | Government hospital, Mission hospital/clinic, Private hospital/clinic | Government health centre, Government dispensary, Other public sector, Nursing/maternity home, Other private sector | Her home, Other home, Other | Doctor | Nurse/midwife | | Traditional Birth Attendant, Community Health Worker, Relative/Friend, Other, No one |
| Liberia | DHS 2007 | Govt. hospital, Private hosp/clinic | Govt. health center, Govt. health clinic, Other public, Other private medical | Respondent's home, Other home, Other | Doctor | Nurse/Midwife | | Traditional midwife, Relative/friend, Other, No one |
| | DHS 2019 | Government hospital, Private hospital/clinic | Govt. health center, Govt. health clinic, Other public, Other private sector | Respondent's home, Other home, Other | Doctor, Physician assistant | Nurse/midwife | | Traditional midwife, Relative/friend, Other, No one |

| | | | | | | | | |
|---|---|---|---|---|---|---|---|---|
| Malawi | DHS 2004 | Govt. hospital, Private hosp/clinic, Mission hospital | Govt. Health Center, Govt. Health Post, Other public, Mission health center, Other private medical | Your home, Other home, Other | Doctor/clinical officer | Nurse/midwife | | Patient attendant, Traditional birth attendant, Relative/friend, Other, No one |
| | MICS 2019 | Government hospital, Private hospital, Cham/mission hospital | Government clinic / health centre, Government health post, Other public, Private clinic, Private maternity home, Other private medical, Cham/mission health centre | Respondent's home, Other home, Other | Doctor/Clinical Officer/ Medical assistant | Nurse/midwife | | Traditional birth attendant, Community health worker/HAS, Relative/friend, Other, No one |
| Mali | MICS 2006 | Hôpital gouvernement | Centre de santé gouv., Poste de santé gouv., Autre public, Hôpital/clinique privée, Matérnite privé, Autre privé médical | Votre domicile, Autre domicile, Autre | Médecin | Sage-femme | | Matrone, Accoucheuse traditionnelle, Parents/AMIS, Autre, Personne |
| | DHS 2018 | Hôpital nat, Hôpital regional | Centre de santé de référence (CSRef), Centre de santé communnautair, Dispensaire/maternite, Autre secteur medical public, Hôpital/clinique privé, Cabinet medical, Cabinet de soin privé, Autre secteur médical privé | Sa maison, Autre maison, Case de santé/asc /relais, Autre | Médecin | Infirmière/sage-femme | Accoucheuse traditionnelle formée/atr | Matrone, Accoucheuse traditionnelle, Agent de santé communaut./ relais, Parent/ami, Autre, Personne |
| Mozambique | DHS 2003 | Hospital do governo, Hospital/clinica privado | Centro/ posto de saúde, Brigadas móveis, Outro sec. Público, Consultorio enfermeira | Sua casa, Casa de parteira tradicional, Casa parteira/ enfermeira, Outro | Médico | Enfermeira/parteira, Parteira auxiliar (auxiliary midwife), | Parteira tradicional (TBA), Atendente | Amigas/FAM., Outre, Ninguém |
| | DHS 2022 | Hospital | Centro de Saúde, Posto de Saúde, Clinica | Própria casa, Outra casa, Outra estabelecimento, Outro privado | Médico | Enfermeira, Parteira (birth attendant) | Parteira tradicional | Amigas/parentes, Outre, Ninguém |
| Niger | DHS 2006 | Hopital govt., Maternite govt., Hôpital/clinique privé | Centre sante, Poste de sante, Autre secteur public, Maternite privé | Votre domicile, Autre domicile, Autre | Médecin | Infirmière/sage femme | Accoucheuse formee | Accoucheuse tradition., Autre, Personne |

| | | | | | | | | |
|---|---|---|---|---|---|---|---|---|
| | DHS 2021 | Hôpital national/reference, Maternité Issaka Gazobi, CHR/ CSME/CHA, Hopital de district, Hôpital/clinique privé/cabinet médical | Centre de santé intégré, Case de santé, Autre secteur public, Pharmacie, Médecin privé | Sa maison, Autre maison, Autre | Médecin | Infirmière/sage femme, Sage-femme auxiliaire | Matronne, Accoucheuse traditionnelle, Agent de sante communautaire/village, Accoucheuse traditionnelle formée | Parent/AMI, Autre, Personne |
| Nigeria | DHS 2003 | Government hospital, Private hospital, | Government health center, Government health station/clinic, Private clinic, NGO Health facility | Respondent's home, Other home, Other | doctor | nurse/midwife | | auxliary midwife, community health worker, traditional birth attendant, relative/friend, other, no one |
| | MICS 2021 | Government hospital, Private hospital | Government health center, Government health post, Other public sector, Private clinic, NGO: health facility, Other NGO health facility | Your home, Other home, Other | doctor | Nurse / Midwife | | Auxilliary Midwife/MCH Aide/Community, Traditional birth attendant, Community health worker, Relative / Friend, Other, No one |
| Rwanda | DHS 2005 | Govt. Hospital, Private hospital/clinic | Govt. health center, other public, other private medical | Your home, other home, other | Doctor | Nurse/ midwife/ medical assistant | Trained traditional birth attendant | Untrained traditional birth attendant, relative/friend, other, no one |
| | DHS 2019 | Ref. hospital, prov./dist. health center | Health center, health post, other public facility, polyclinic, clinic, dispensary, other private med. facility | Her home, other home, other | Doctor | Nurse/ midwife | Auxiliary nurse midwife, | Traditional birth attendant, community health worker, community health mother and child, relative/friend, other, no one |
| Senegal | DHS 2005 | hopital gouvt, hopital/clinique prive, | centre de sante/maternite, Poste de santé Gouvt, autre public, autre prive medical | Votre domicile, Autre domicile, Autre | Médecin | Sage-femme, Infirmiére/ICP | | Matronne, Accoucheuse traditionnelle, Parent, amie, autre (spécifique au pays), Autre, Personne |
| | DHS 2019 | Hôpital Gouvt, Hôpital/Clinique privé | Centre de santé/maternité, Poste de santé Gouvt, Case de santé, Clinique mobile, Autre secteur public, Autre secteur médical privé | Sa maison, Autre maison, Pharmacie, Boutique, Praticien traditionne, Marché agent, Autre | Médecin | Sage-femme, Infirmiére/ICP | | Matronne, Accoucheuse traditionnelle, Autre, Personne |

| | | | | | | | | |
|---|---|---|---|---|---|---|---|---|
| Sierra Leone | MICS 2005 | Govt. hospital, Private hospital | Govt. clinic/health center, Other public, Private clinic, Private maternity home, Other private medical | You home, Other home, Other | Doctor | Nurse/midwife | Auxiliary midwife/ MCH Aide, | Traditional Birth Attendant, Community Health Worker, Relative/friend, Other, No one |
| | DHS 2019 | Government hospital, Private hospital | Government clinic / health centre, Government health post, Other public, Private clinic, Private maternity home | Her home, Other home, Other | Doctor | Nurse/midwife | Other qualified?, MCH Aide (not in questionnaire) | Traditional birth attendant, Community/Village health worker, Relative / Friend, Other, No one |
| Tanzania | DHS 2004 | Referal/ spec. hospital, Regional hospital, District hospital, Referal/spec. hospital, District hospital, Religious: referal/spec. hospital, Religious: district hospital | Health centre, Dispensary, Village health post, Govt. health centre, Religious: health centre, Religious: dispensary (Cbd worker) | Respondent's home, Other home, Other | Doctor | Nurse/Midwife | Clinical Officer, Trained birth attendant | Auxilliary Midwife/MCH Aide, Assistant Clinical Officer, Traditional birth attendant, Relative/Friend, Village health worker, Other, No one |
| | DHS 2022 | National/zonal referral/ spec. hospital, Regional referral hospital(government/parastatal), Regional hospital(government/parastatal), District hospital (government/parastatal), Referal/ spec. hospital (religious/voluntary), District hospital (religious/voluntary), Hospital (religious/voluntary), Clinic (religious/voluntary), Specialized hospital (private) | Health center (government/parastatal), Dispensary (government/parastatal), Clinic (government/parastatal), Health center (religious/voluntary), Dispensary (religious/voluntary), Hospital (private), Health centre (private), Dispensary(private) | Respondent's home, Other home, TBA premises, Other | Doctor/amo | Nurse/midwife, Assistant nurse | Clinical officer | Assistant clinical officer, MCH AIDE, CHW, Other, Traditional Birth Attendant, Relative/friend, No one |
| Uganda | DHS 2006 | GOVT. HOSPITAL, PVT. HOSPITAL/CLINIC | GOVT. HEALTH CENTER, GOVT. HEALTH POST, OTHER PUBLIC, OTHER PRIVATE MED. | YOUR HOME, TBA's HOME, OTHER HOME, OTHER | Doctor | Midwife/ Nurse | Medical Assistant, Clinical Officer, Nursing Aide | Traditional Birth Attendant, Relative/Friend, Other, No one |
| | DHS 2016 | Government hospital, Private hospital/clinic | Government health center, Other public | Her home, Other | Doctor | Midwife/ Nurse | Medical assistant/clinical officer, | Traditional Birth Attendant, |

| | | | | | | | | |
|---|---|---|---|---|---|---|---|---|
| | | | sector, Other private medical sector | home, Other | | | Nursing aide/assistant | Relative/Friend, Other, No one |
| Zambia | DHS 2001 | Govt. Hospital, Pvt. Hospital/clinic, Mission hospital/clinic | Govt. Health post, Govt. Health center, | Your home, Other home, Other | Doctor | Nurse/midwife | Clinical officer | Traditional Birth Attendant, Relative/Friend, Other, No one |
| | DHS 2018 | Government hospital, Private hospital/clinic, Mission hospital/clinic | Government health post, Government health center, Other public sector, Other private medical sector | Her home, Other home, Other | Doctor | Nurse/midwife | Clinical officer | Traditional Birth Attendant, Community/village healht assistant, Relative/Friend, Community/village health worker, Other, No one |
| Zimbabwe | DHS 2005 | Central hsp, Provincial hsp, Dist/rural hsp, Private hsp/clc. | Rural/muncpl cl, Rural hlth cntr, Other public | Your home, Other home, Other | Doctor | Nurse midwife | Trained birth attendant | Untrained (traditional birth attendant), unsure about training, Other, No one |
| | MICS 2019 | Government hospital, Government Clinic / Health Centre, Private hospital, Mission hospital | Council facility, Private clinic, Mission clinic, Other mission | Respondent's home, Other home, Other | Doctor | Nurse/midwife | | Traditional Birth Attendant, Community Health Worker, Relative / Friend, Religious birth attendant, Other, No one |

#### Table A.3: HMIS facility level categorizations

| | **Hospital Categories** | **Non-Hospital Categories** |
|---|---|---|
| **Burkina Faso** | Regional Hospital (CHR), Medical Centre with a Surgical Unit (CMA), University Teaching Hospital (CHA) | Clinique, Medical Centre (CM), Health and Social Promotion Centre (CSPS), Dispensaire, Infirmerie, Cabinet, CREN, Maternités Isolées |
| **Cameroon** | Hospital, Clinic | Dispensaire, Health Centre |
| **Cote d'Ivoire** | Centre Hospitalier Regional, Hopital General, Hopital | Centre De Sante, Formation Sanitaire, Infirmerie, Polyclinique/Clinique, Maternité, Centre Médical Spécialisé (Specialized Medical Center) (CMS), Centre De Santé Urbain (Urban Health Center) (CSU) |
| **DRC** | Hospital | Health Center, Health Station, Health Posts, Medical Center, Polyclinique, Clinics, Dispensary, Maternity |
| **Ethiopia** | Hospitals | Health centers, clinics, health posts, specialty centers |
| **Ghana** | Hospital, District Hospital, Teaching Hospital, Regional Hospital, University Hospital/Clinic, Psychiatric Hospital | Community Health and Planning Services (CHPS), Clinic, Health Centre, Maternity Home, Polyclinic, Medical Centre |
| **Guinea** | Hopital, Centre Medical Communal (CMC) | Centre De Santé, Infirmerie, Institut, Dispensaire, Centre De Santé Amélioré, Cabinets de Soin |
| **Kenya** | Hospitals | Basic Health Centre, Dispensary, Medical Clinic, Health Centre, Comprehensive Health Centre, Medical Center, Nursing and Maternity Home, Nursing Homes, Nursing Home, Radiology Clinic, VCT |
| **Liberia** | Hospital | Clinic, Health Center |
| **Malawi** | Hospital | Health Centre, Clinic, Dispensary, Maternity |
| **Mali** | Hôpital | Dispensaire, Maternité |

| | | |
|---|---|---|
| **Mozambique** | Hospital | Clinic |
| **Niger** | Hospital, Centre Hospitalier Régional (CHR) | Dispensary, Maternite |
| **Nigeria** | Hospital, State Hospital, Federal Teaching Hospital, Specialist Hospital, University Teaching Hospital, | Maternity/Nursing Home, Clinic, Health Centre/Post/Care/Clinic, Medical Centre, Dispensary, Infirmary, Specialist |
| **Rwanda** | Private Hospital, District Hospital (DH), Referral Hospitals (RH), Provincial Hospital (PH) | Health Center (CS), Private Clinic, Health Post (HP) |
| **Senegal** | EPS | CS, PS, Clinique, Cabinet |
| **Sierra Leone** | Hospital | Community Health Center (CHC), Community Health Post (CHP), Clinic, Maternal and Community Health Post (MCHP), outreach |
| **Tanzania** | Hospital | Clinic, Dispensary, Health Centre |
| **Uganda** | Hospital, Regional Referral Hospital (RRH), National Referral Hospital (NRH) | Clinic, Nursing Home, Health Centre, Medical Centre, Domiciliary, Community Centre, Maternity |
| **Zambia** | Hospital | Clinic, health centre, health post |
| **Zimbabwe** | Hospital | Clinic, Health Post, Health Centre |

#### Table A.4: Country HMIS 2022 MNH data reporting

| Countries | Total facilities in HMIS 2022 reporting MNH data | Proportion of hospitals (%) | Merging rate between MNH and Administrative records (%) | Proportion of hospitals reporting any CS (%) | Proportion of non-hospitals reporting any CS (%) |
|---|---|---|---|---|---|
| **Burkina Faso** | 2,512 | 2.6 | 99.9 | 95.5 | 1.2 |
| **Cameroon** | 5,293 | 2.2 | 98.3 | 78.4 | 13.1 |
| **Cote d'Ivoire** | 2,692 | 4.9 | 96.7 | 57.1 | 0.2 |
| **DRC** | 18,155 | 6.2 | 96.9 | 89.2 | 6.7 |
| **Ethiopia** | 6,919 | 5.9 | 93.1 | 93.9 | 2.7 |
| **Ghana** | 4,350 | 13.4 | 99.5 | 42.7 | 0.7 |
| **Guinea** | 515 | 6.8 | 47.8 | 100.0 | 4.2 |
| **Kenya** | 5,812 | 13.8 | 89.7 | 0.6 | 0.1 |
| **Liberia** | 733 | 5.0 | 99.7 | 83.8 | 2.3 |
| **Malawi** | 599 | 15.2 | 97.7 | 74.7 | 14.0 |
| **Mali** | 1,745 | 4.4 | 100.0 | 97.4 | 0.1 |
| **Mozambique** | 1,584 | 4.3 | 100.0 | 91.2 | 1.7 |
| **Niger** | 3,216 | 1.4 | 100.0 | 82.2 | 0.1 |
| **Nigeria** | 20,995 | 11.7 | 53.7 | 75.6 | 9.5 |
| **Rwanda** | 609 | 8.9 | 100.0 | 98.1 | 1.8 |
| **Senegal** | 1,809 | 1.9 | 100.0 | 94.1 | 2.9 |
| **Sierra Leone** | 1,408 | 1.8 | 99.9 | 88.5 | 6.9 |
| **Tanzania** | 6,709 | 0.9 | 98.3 | 100.0 | 11.6 |
| **Uganda** | 4,000 | 4.8 | 94.0 | 85.4 | 8.2 |
| **Zambia** | 2,587 | 6.7 | 100.0 | 84.4 | 2.3 |
| **Zimbabwe** | 1,537 | 12.4 | 98.4 | 44.0 | 0.4 |

**Note:** For reporting completeness, if a facility did not have all 12 monthly reports, adjustments were made by multiplying the yearly volume by the reciprocal of the reporting rate to impute the missing monthly data point. Extreme outliers were identified using the Hampel X84 method, using a modified Z-score as a standardised score of data points measuring the deviation from the median, obtained by dividing the difference from the median by the median absolute deviation. Monthly data with a score greater than five standard deviations from the annual median were considered extreme outliers. We adjusted any extreme outliers and missing data with the monthly median for the year.

#### Appendix B

##### Table B.1: Country socio-demographic and mortality data, UN estimates 2022-3

| Countries | Population in 2022, millions [55] | Total births in 2022, millions [55] | Total fertility rate in 2022, children per woman [55] | Neonatal mortality rate in 2023, per 1000 live births [1] | Stillbirth rate in 2023, per 1000 births [1] | Maternal mortality ratio in 2023, per 100,000 live births (80% uncertainty interval) [2] | Mortality transition phase*, 2023 |
|---|---|---|---|---|---|---|---|
| **Burkina Faso** | 22.5 | 0.74 | 4.27 | 24 (14-41) | 19 (17-21) | 242 (155-367) | 3 |
| **Cameroon** | 27.6 | 0.98 | 4.40 | 25 (18-35) | 18 (11-31) | 258 (196-359) | 3 |
| **Cote d'Ivoire** | 30.39 | 1.02 | 4.35 | 28 (22-35) | 23 (16-32) | 359 (237-568) | 3 |
| **DRC** | 102.39 | 4.56 | 6.11 | 25 (13-46) | 26 (21-32) | 427 (283-775) | 3 |
| **Ethiopia** | 125.38 | 4.18 | 4.08 | 27 (20-38) | 30 (24-37) | 195 (128-332) | 3 |
| **Ghana** | 33.15 | 0.90 | 3.43 | 21 (17-26) | 19 (14-26) | 234 (155-344) | 3 |
| **Guinea** | 14.06 | 0.50 | 4.30 | 30 (21-43) | 23 (14-38) | 494 (337-764) | 2 |
| **Kenya** | 54.25 | 1.54 | 3.26 | 22 (17-26) | 16 (15-17) | 379 (267-547) | 3 |
| **Liberia** | 5.37 | 0.17 | 4.02 | 30 (20-44) | 24 (14-42) | 628 (436-913) | 2 |
| **Malawi** | 20.57 | 0.69 | 3.72 | 19 (11-33) | 16 (12-20) | 225 (153-352) | 3 |
| **Mali** | 23.07 | 0.99 | 5.69 | 32 (23-46) | 23 (15-34) | 367 (268-498) | 2 |
| **Mozambique** | 32.66 | 1.30 | 4.84 | 25 (20-33) | 18 (14-23) | 82 (60-113) | 3 |
| **Niger** | 25.31 | 1.14 | 6.14 | 34 (25-47) | 21 (14-33) | 350 (227-563) | 3 |
| **Nigeria** | 223.15 | 7.64 | 4.55 | 34 (23-51) | 24 (14-40) | 993 (718-1540) | 1/2 |
| **Rwanda** | 13.65 | 0.40 | 3.78 | 18 (13-24) | 16 (12-22) | 229 (158-373) | 3 |
| **Senegal** | 17.65 | 0.55 | 3.86 | 22 (18-28) | 18 (15-22) | 237 (173-365) | 3 |
| **Sierra Leone** | 8.28 | 0.26 | 3.88 | 29 (22-39) | 20 (12-35) | 354 (249-537) | 3 |
| **Tanzania** | 64.71 | 1.74 | 4.67 | 21 (16-27) | 19 (14-26) | 276 (192-429) | 3 |
| **Uganda** | 47.31 | 2.42 | 4.39 | 18 (11-28) | 15 (12-17) | 170 (116-298) | 3 |
| **Zambia** | 20.15 | 0.71 | 4.17 | 22 (14-33) | 14 (12-16) | 85 (61-126) | 3 |
| **Zimbabwe** | 16.07 | 0.50 | 3.77 | 22 (13-37) | 19 (17-23) | 358 (236-484) | 3 |

*Phase 2 and 3 are marked by an MMR of 300-700 and 100-300 per 100,000 live births respectively, and stillbirth rate (SBR) plus neonatal mortality rate (NMR) of 55-80 and 30-55 per 1000 births respectively (30-45 and 15-30 per 1000 for NMR alone). The latter corresponds with neonatal mortality less than 15 per 1,000 live births [5].

##### Table B.2: Transition model typical values for coverage indicators [5]

| COVERAGE INTERVENTIONS | PHASE | MEDIAN | 25TH PERCENTILE | 75TH PERCENTILE |
|---|---|---|---|---|
| Institutional delivery (%) | I | 35.6 | 24.6 | 48.2 |
| | II | 59.4 | 42.6 | 74.9 |
| | III | 73.6 | 57.2 | 90.2 |
| | IV | 96.9 | 87.7 | 98.6 |
| | V | 99.3 | 98.6 | 99.8 |
| Institutional delivery, poorest quintile (%) | I | 13.6 | 9.5 | 24.8 |
| | II | 35.2 | 18.5 | 54.5 |
| | III | 45.4 | 26.4 | 72.5 |
| | IV | 91.8 | 69.0 | 96.9 |
| | V | 99.1 | 97.5 | 99.7 |
| Institutional delivery, absolute gap between richest and poorest quintile (percentage points) | I | 53.9 | 36.0 | 61.3 |
| | II | 51.0 | 37.8 | 61.9 |
| | III | 44.1 | 20.9 | 62.6 |
| | IV | 7.8 | 1.2 | 28.9 |
| | V | 0.2 | -0.3 | 1.0 |
| Hospital delivery (%) | I | 18.6 | 11.2 | 28.3 |

| | | | | |
|---|---|---|---|---|
| | II | 26.3 | 15.8 | 33.9 |
| | III | 48.2 | 28.1 | 67.8 |
| | IV | 72.8 | 57.0 | 94.4 |
| | V | 97.6 | 92.3 | 98.7 |
| Lower-level facility delivery (%) | I | 12.7 | 6.0 | 17.6 |
| | II | 29.6 | 12.7 | 44.3 |
| | III | 15.7 | 3.1 | 31.2 |
| | IV | 17.7 | 3.0 | 30.9 |
| | V | 2.3 | 0.8 | 6.4 |
| C-section rate (%) | I | 2.3 | 1.5 | 4.2 |
| | II | 4.5 | 3.0 | 6.4 |
| | III | 12.9 | 6.3 | 21.7 |
| | IV | 25.8 | 18.6 | 33.2 |
| | V | 25.3 | 18.4 | 29.6 |
| C-section rate, poorest quintile (%) | I | 0.8 | 0.3 | 1.7 |
| | II | 1.7 | 0.9 | 2.7 |
| | III | 4.3 | 2.1 | 10.7 |
| | IV | 15.1 | 7.4 | 21.9 |
| | V | 18.2 | 14.2 | 29.0 |

#### Appendix C

**Table C.1: C-Section rates among live births in the three years preceding, overall and among the poorest and richest groups in early and recent surveys (DHS/MICS)**

| Delivery coverage group in recent survey | Country | Survey years | C-section rate (per 100 live births in the three years preceding)** | | | | | |
|---|---|---|---|---|---|---|---|---|
| | | | Overall | | Poorest (Q1) | | Richest (Q5) | |
| | | | Early | Recent | Early | Recent | Early | Recent |
| Lowest (<65%) | Niger | 2006-2021 | 1.0 | 1.8 | 0.2 | 1.2 | 3.9 | 5.6 |
| | Ethiopia | 2005-2019 | 1.1 | 5.7 | 0.1 | 1.6 | 6.1 | 12.9 |
| | Nigeria | 2003-2021 | 1.7 | 3.8 | 0.6 | 1.3 | 4.8 | 10.6 |
| | Guinea | 2005-2018 | 1.8 | 2.9 | 0.2 | 0.4 | 7.5 | 7.3 |
| Medium | Mozambique | 2003-2022 | 2.2 | 5.1 | 0.4 | 1.2 | 9.3 | 14.5 |
| | Mali | 2006-2018 | 1.7 | 2.5 | 1.1 | 1.7 | 5.0 | 6.0 |
| | Cameroon | 2004-2018 | 2.1 | 4.1 | 0.6 | 0.9 | 4.5 | 11.2 |
| | Uganda | 2006-2016 | 3.4 | 6.7 | 2.2 | 2.6 | 8.2 | 16.4 |
| | Burkina Faso | 2003-2021 | 0.7 | 6.3 | 0.2 | 4.8 | 2.2 | 12.1 |
| | Tanzania | 2004-2022 | 3.6 | 10.5 | 1.1 | 4.1 | 9.3 | 22.3 |
| | Liberia | 2007-2019 | 4.0 | 5.7 | 0.9 | 2.5 | 8.3 | 10.5 |
| | Senegal | 2005-2019 | 3.6 | 7.9 | 0.7 | 2.6 | 8.1 | 17.6 |
| | Cote d'Ivoire | 2011-2021* | 3.1 | 8.5 | 0.5 | 2.8 | 7.9 | 19.4 |
| | DRC | 2007-2017 | 4.5 | 4.7 | 2.7 | 1.9 | 6.3 | 10.5 |
| | Sierra Leone | 2008-2019* | 1.8 | 4.9 | 0.6 | 3.3 | 6.1 | 10.4 |
| | Zambia | 2001-2018 | 2.2 | 5.5 | 0.4 | 2.7 | 7.1 | 14.4 |
| Highest (>85%) | Ghana | 2003-2022 | 4.2 | 20.3 | 1.5 | 11.1 | 13.3 | 37.7 |
| | Zimbabwe | 2005-2019 | 4.9 | 8.8 | 1.4 | 4.5 | 11.4 | 5.2 |
| | Kenya | 2003-2022 | 4.3 | 16.8 | 1.2 | 5.5 | 11.6 | 31.7 |
| | Rwanda | 2005-2019 | 3.0 | 16.2 | 1.2 | 11.4 | 8.3 | 29.8 |
| | Malawi | 2004-2019 | 3.2 | 7.7 | 3.6 | 4.9 | 4.8 | 16.1 |
| | Median | 2005-2019 | 3.0 | 5.7 | 0.7 | 2.6 | 7.5 | 12.9 |

*Early surveys did not report C-section rates so the next subsequent survey estimates were used.

** Some estimates may differ slightly from survey reports because the years preceding the survey led to different denominators of live births used for the estimate.

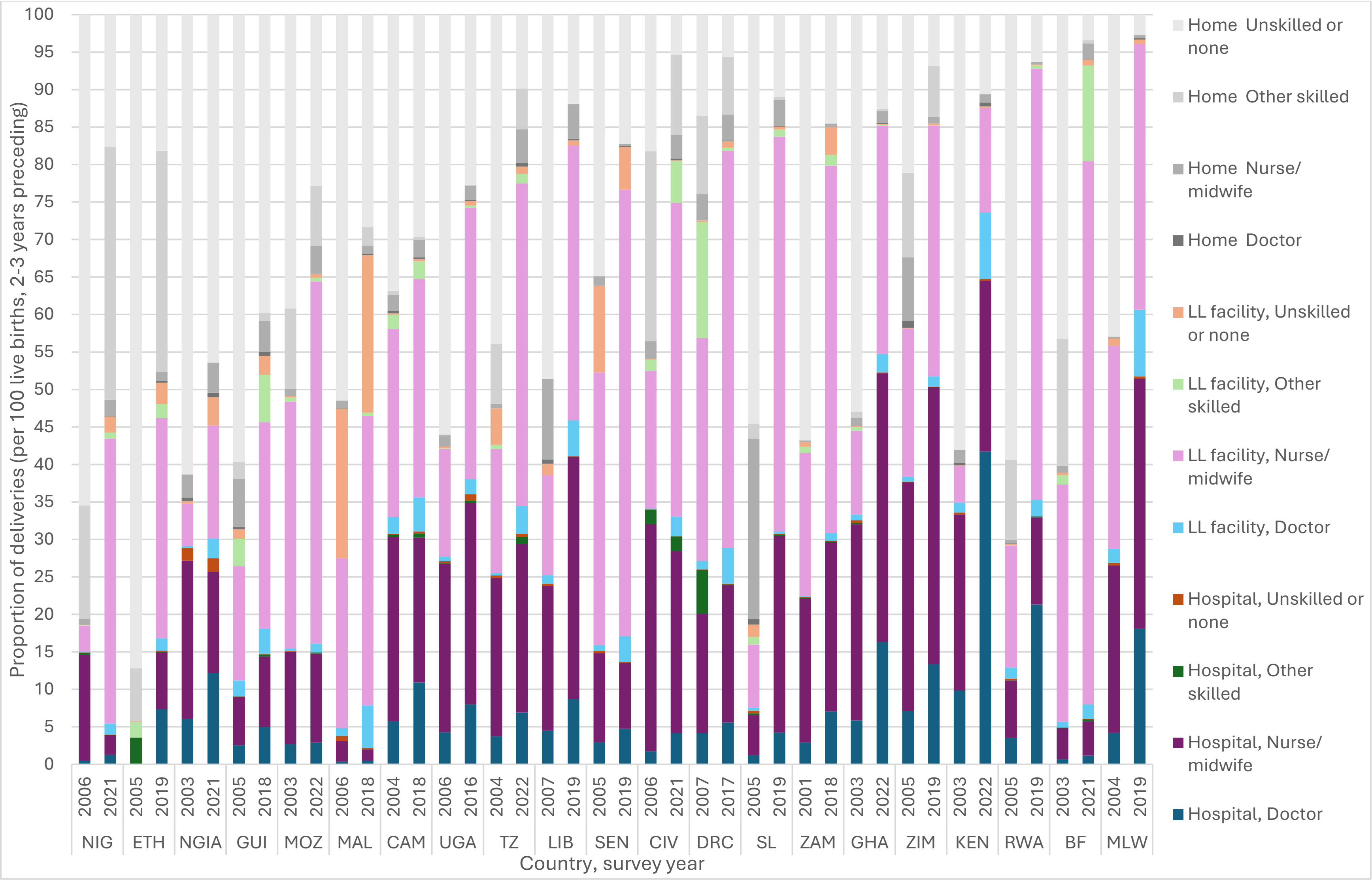


**Figure C.1: Trends in the proportion of deliveries by facility level and birth attendant type (early and recent DHS/MICS)**

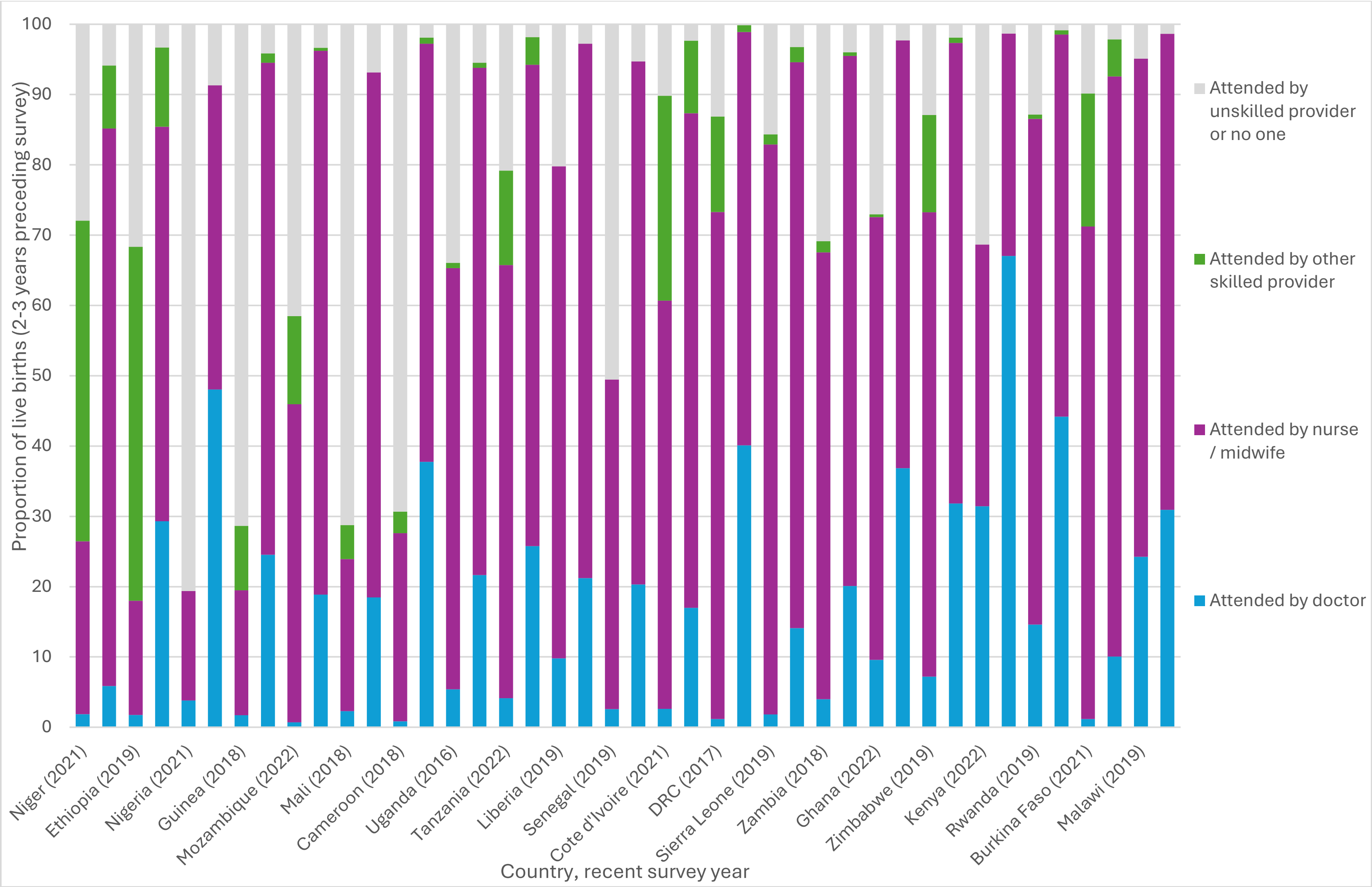


**Figure C.2: Proportion of deliveries by attendant type among the poorest (Q1) and richest (Q5) groups (recent DHS/MICS)**

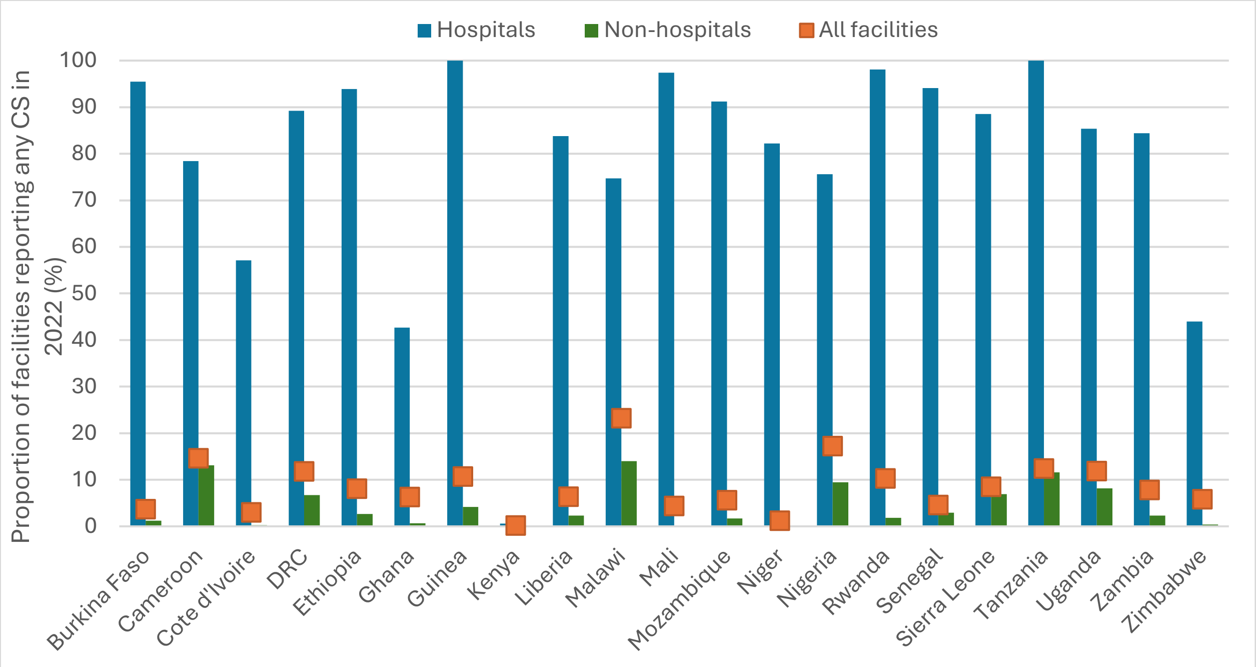


**Figure C.3: Proportion of hospitals and non-hospitals reporting any C-sections (HMIS 2022)**

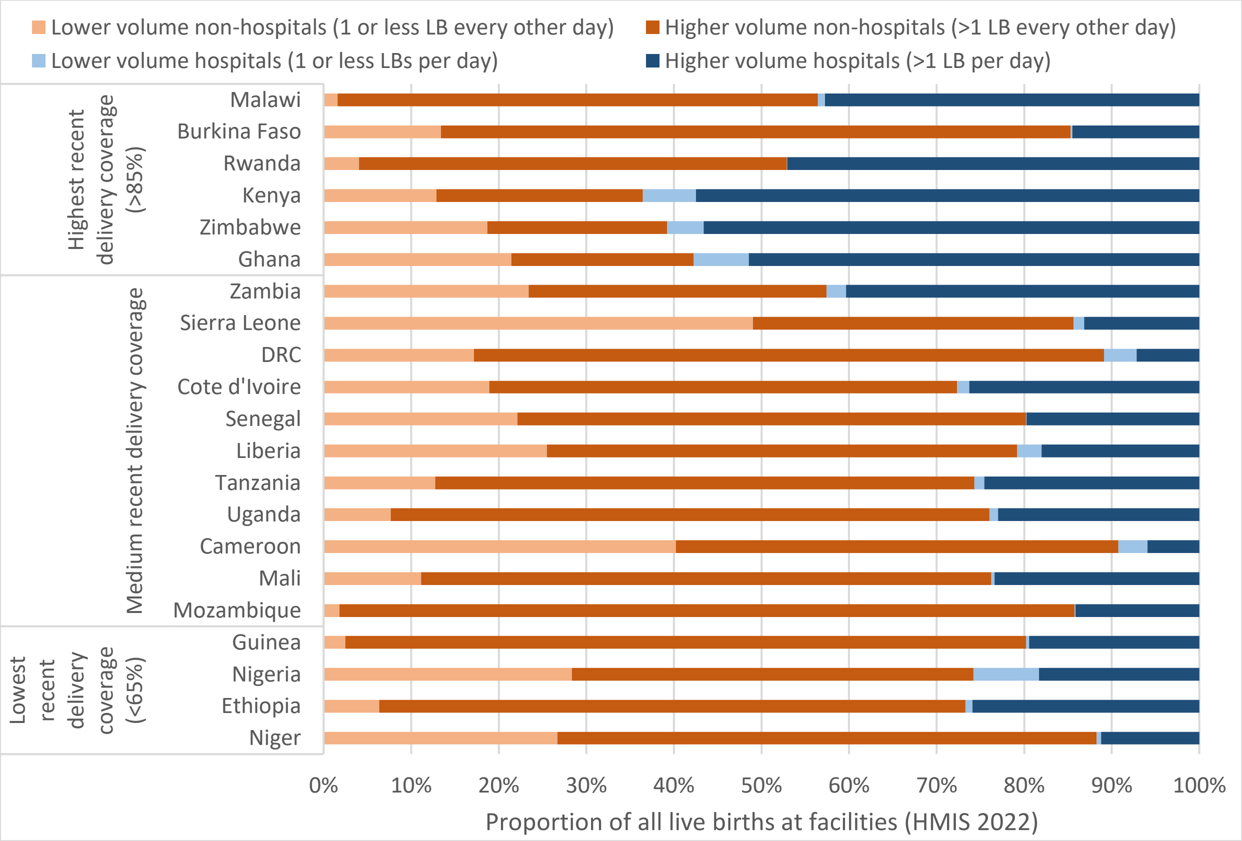


**Figure C.4: Proportion of all live births at facilities with lower and higher median live birth volumes at non-hospitals and hospitals in 2022, by recent delivery coverage group (HMIS)**

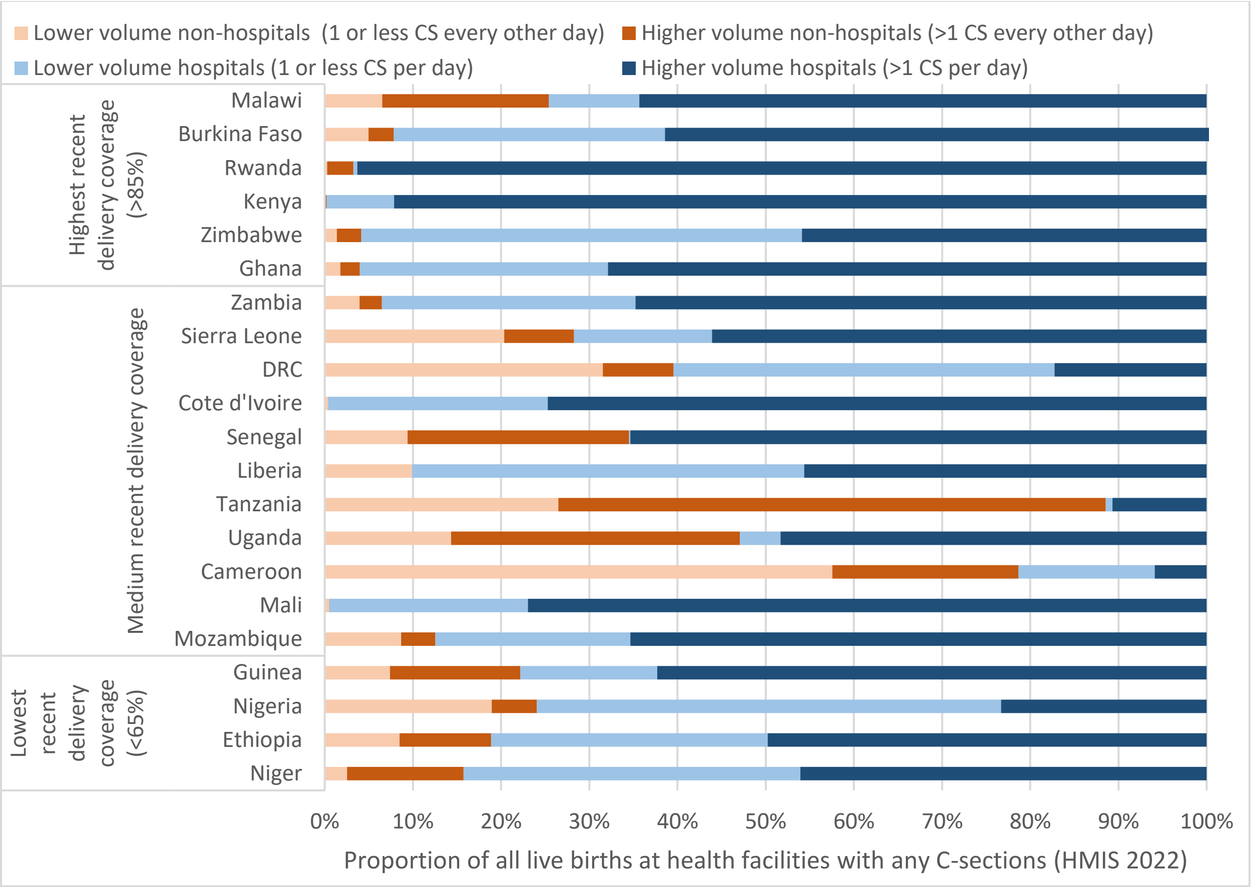


**Figure C.5: Proportion of all live births at facilities with lower and higher median C-section (CS) volumes at non-hospitals and hospitals conducting any CS in 2022, by recent delivery coverage group (HMIS)**
